# Sequencing individual genomes with recurrent genomic disorder deletions: an approach to characterize genes for autosomal recessive rare disease traits

**DOI:** 10.1101/2021.02.16.21251842

**Authors:** Bo Yuan, Katharina Schulze, Nurit Assia Batzir, Jefferson Sinson, Hongzheng Dai, Wenmiao Zhu, Francia Bocanegra, Chin-To Fong, Jimmy Holder, Joanne Nguyen, Christian P. Schaaf, Yaping Yang, Weimin Bi, Christine Eng, Chad Shaw, James R. Lupski, Pengfei Liu

## Abstract

In medical genetics, discovery and characterization of disease trait contributory genes and alleles depends on genetic reasoning, study design, and patient ascertainment; we now suggest a segmental haploid genetics approach to enhance gene discovery and molecular diagnostics. We present novel genomic insights to enhance discovery in the challenging context of autosomal recessive (AR) traits and bi-allelic variation. We demonstrate computationally that new mutation mediated by nonallelic homologous recombination (NAHR), involving recurrent deletions at 30 genomic regions, likely drives recessive disease burden for over 70% of loci within these segmental deletions or at least 2% of loci genome wide. Meta-analyses of literature-reported patients implicate that NAHR-deletions are depleted from the ascertained pool of AR trait alleles. Exome reanalysis of personal genomes from subjects harboring recurrent deletions uncovered new disease-contributing variants in genes including *COX10*, *ERCC6, PRRT2* and *OTUD7A*. Our data demonstrate that genomic sequencing of personal genomes with NAHR-deletions could dramatically foment allele and gene discovery, enhance clinical molecular diagnosis, and could potentially enable human haploid genetics screens as an approach to disease biology.

## Introduction

During the previous decade, and since the first personal genome by massively parallel DNA sequencing of the JDW genome (Wheeler et al. 2008), efforts to decipher molecular and genetic mechanisms underlying Mendelian conditions have repeatedly demonstrated that mutations aggregate in personal genomes and can cause human diseases in a continuum of allelic modalities, ranging from monoallelic (dominant), biallelic (recessive), triallelic, to multiallelic and more complex modes of inheritance (Lupski et al. 2011; Lupski 2021). Recent large-scale family-based genomic studies using exome sequencing (ES) have uncovered hundreds of new disease loci, with the majority following traditional Mendelian inheritance, i.e., monoallelic (autosomal dominant [AD]) or biallelic (autosomal recessive [AR]) trait segregation (Posey et al. 2019; Baxter et al. 2022). Although optimism has been increasing towards achieving disease annotation for a substantial portion of the haploinsufficient part of the human genome through dominant disease gene discoveries, statistical analysis from rare disease cohort studies suggests that the trajectory to understanding, or illumination of the biology thereof, of the rare recessive disease traits, that is specifically the biallelic-disease-causing portions of the human genome, is less certain (Martin et al. 2018; Bamshad et al. 2019; Cacheiro et al. 2020).

This apparent discrepancy in discovery between AD and AR trait genes can perhaps most parsimoniously be explained by the Clan Genomics Model (Lupski et al. 2011). The model predicts that dominant diseases are largely caused by emergence of new alleles in recent generations, *i.e. de novo* mutations (DNMs). On the other hand, recessive disease traits arise when a *pair* of disease alleles at a locus are aggregated within a personal genome in the *trans* configuration by transmission genetics, i.e. maternal + paternal haplotypes; the incidence of disease correlates with the product of the probabilities of sampling each of the two alleles, whether existing in a population or *de novo*, in a mating. Because of the bivalent nature of such alleles, the overall incidence of the recessive trait depends on the local allelic architecture, i.e., the snapshot of the frequency distribution of all pathogenic alleles. This is a characteristic not observed in dominant disease traits, wherein the individual alleles act in solitude and the overall incidence mostly depends on new mutations.

Individual recessive trait alleles can emerge and be carried in populations, clans and pedigrees with no impact on phenotype. Consequently, some recessive disease alleles can reach high population allele frequencies [e.g., 8×10^-4^ for NM_000520.6(*HEXA*):c.1274_1277dup (p.Tyr427fs), the most common Tay Sachs disease allele], which can be several orders of magnitude higher than alleles associated with dominant disease traits (∼10^-8^ as an estimate for the *de novo* single nucleotide variant rate). As one may surmise or expect, previous research efforts or clinical testing in unselected cohorts are likely to ascertain recessive trait diseases with at least one allele of high population frequency. In contrast, if a yet-to-be-defined recessive disease trait gene does not have appreciable pathogenic alleles represented at a sufficient population frequency, disease discovery and annotation of the gene would be greatly hampered due to the extremely low incidence and difficulty in ascertaining affected individuals, even when considering a world-wide population of 7.8 billion.

Special strategies and genetic and genomic approaches need to be implemented to overcome this potential ‘barrier to discovery’ and characterization initiatives. Ascertaining patients in populations with an elevated coefficient of consanguinity and autozygosity is a widely applied and highly successful strategy for rare recessive trait disease gene discovery. In this circumstance, the allele pool shrinks to the Clan of the patient’s extended family, which dramatically escalates the effective disease-allele frequency compared to the baseline allele frequency in the general population (Gonzaga-Jauregui et al. 2020; Alkuraya 2021). Thus, the probability of ascertaining patients with a recessive trait disorder increases exponentially, because the sampling of the second allele occurs within the Clan rather than the general population (Coban-Akdemir et al. 2020). Similarly, focusing on a specific geographic or ethnic population is another effective strategy, often attributed to available founder mutation alleles in the population studied (Gonzaga-Jauregui et al. 2020); the Puerto Rican founder allele, and specifically the *COL27A1* founder in a geographic isolation bottleneck and Steel syndrome on the island of Puerto Rico, was informative. The genetic and genomic data from the United States and in populations from countries mapping to multiple continents provide further support to the Clan Genomics hypothesis worldwide (Gonzaga-Jauregui et al. 2020).

Here, we present an alternative study design strategy to enhance the investigation of novel AR disease trait genes, and alleles in biallelic recessive traits, that are difficult to access by conventional methods. This strategy leverages loss-of-function (LoF) alleles caused by large recurrent genomic deletions rendering a locus haploinsufficient (for AD traits) or hemizygous (for AR traits). Recurrent genomic deletions are a subset of contiguous gene deletions that are characterized by a special type of mutational mechanism called nonallelic homologous recombination, or NAHR (Lupski 2019). NAHR is mediated by ectopic recombination between highly similar repeat sequences termed low-copy repeats (LCRs) or segmental duplications (SDs) (Liu et al. 2012). The human genome is evolutionarily structured to be highly enriched for SDs, which creates a large number of architectural hotspots for genomic disorders and mirror traits to emerge (Lupski 2015b; Dennis and Eichler 2016).

Recurrent genomic deletions (or NAHR-deletions) set up an ideal background for AR disease trait gene discovery due to a key property: they act as highly prevalent recessive alleles in comparison to other small variant, i.e. single nucleotide variants (SNV) and indels, recessive disease alleles and they continue to renew because of high mutation rates of structural variant mutagenesis. It has been shown that the mutation rate of NAHR at a given locus can be as high as ∼10^-4^ to 10^-5^, which is orders of magnitudes higher than the per base new mutation rates from SNVs and indels (Turner et al. 2008). The high new mutation rate ensures that these genomic deletions continuously arise *de novo* in the human population among unrelated individuals (Berg et al. 2010; Beck et al. 2019). This ‘recurrent’ nature of genomic deletions distinguishes them from the other recessive trait disease alleles that are more ‘stationary’ or ancestral artifacts of past population history amplified by recent population expansion. Moreover, as genotyping assays are performed on relatives of patients with deletions as well as on individuals without a disease indication, we recognize that many recurrent genomic deletions are incompletely penetrant (Potocki et al. 1999; Rosenfeld et al. 2013; Stefansson et al. 2014; Wu et al. 2015; Crawford et al. 2019; Yang et al. 2019; Collins et al. 2020; Ren et al. 2020), i.e., the fitness of the deletion allele can be high, at least in certain genomic backgrounds (Lupski 2015a; Mannik et al. 2015).

Thus, we hypothesized that because of these key attributes, population prevalence and new mutation rate, NAHR alleles are contributing to a considerable fraction of recessive rare diseases traits at loci mapped within NAHR genomic intervals and may be among the most relevant and prominent alleles at these recessive trait loci. Additionally, we hypothesize that although the sequencing of large recurrent deletions has resulted in isolated characterizations of new recessive genes and alleles in the past, such focused efforts have been under-recognized as a concerted generalizable study strategy, possibly due to the preconceived notion that most of the large recurrent deletions are ‘dominant’ incompletely penetrant disease alleles.

To pursue these hypotheses, we formulated a large computational analysis using both genome-wide population data resources as well as clinical genomics empirically derived information (Figure 1). Herein, we demonstrate, using computational and sequencing analyses, that recurrent NAHR-deletions contribute to a major fraction of individual disease burden for over 2% of known recessive trait genes or 70% of known recessive disease traits in regions encompassed by LCRs; the latter genomic intervals known to undergo NAHR at elevated mutational rates (Turner et al. 2008; Berg et al. 2010) (Figure 2). By meta-analysis of all patients and disease alleles reported in the literature from the top rare recessive disease trait genes predicted to be driven by NAHR-deletion alleles, we present evidence suggesting that the genomic sequencing of individuals with recurrent deletions is under-utilized. The findings regarding allelic architecture of diseases, leveraging new mutation and the high incidence of recurrent genomic deletion alleles, can ‘prime’ powerful future strategies for recessive disease trait genes and allele discoveries exploiting the concept of ‘human haploid genetics’, from the original utilization of disease trait genes mapping on the X chromosome in affected males (Ballabio and Andria 1992), to the current proposed application of investigating genomic intervals of recurrent segmental aneusomy for each of the diploid autosomal chromosomes.

**Figure 1.**
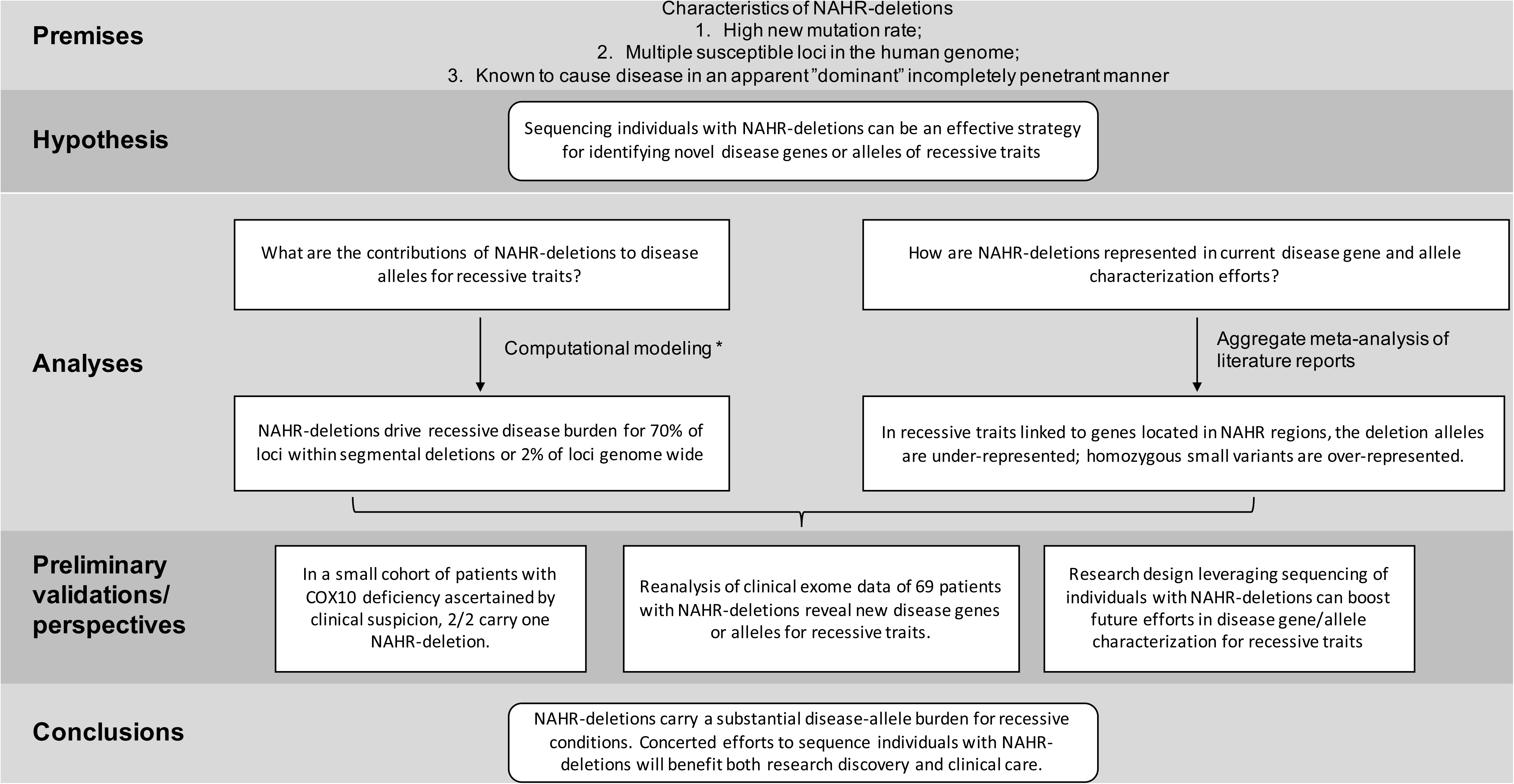
A flowchart of this study. Abbreviations: NAHR, non-allelic homologous recombination. *, a summary of the computational modeling is provided in Figure 2.

**Figure 2.**
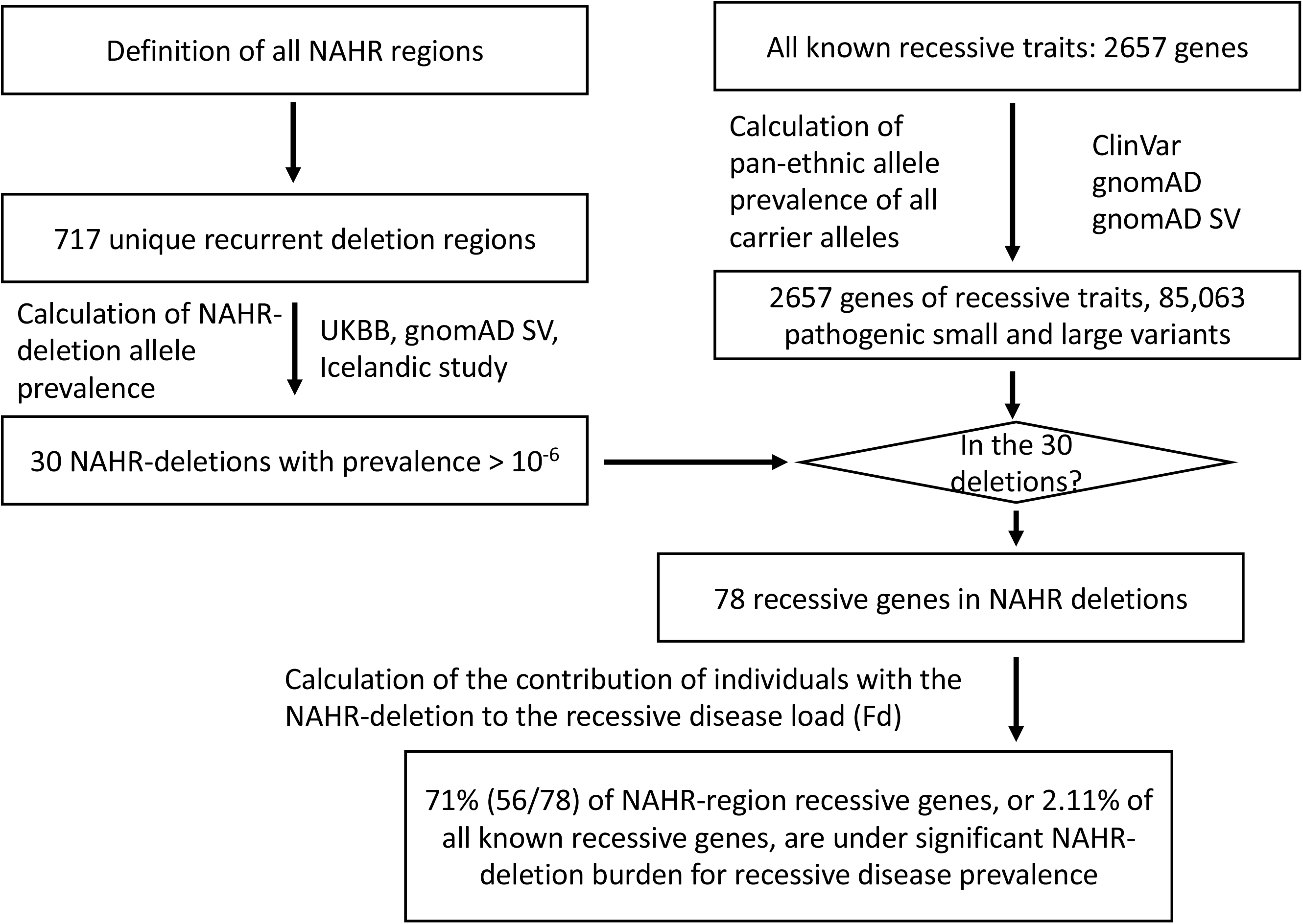
Computational modeling reveals unexpected contribution of NAHR-deletions to autosomal recessive disease trait burden.

## Results

### NAHR-deletion: the most prevalent disease allele for a major fraction of recessive trait genes mapping to 30 genomic loci

In order to systematically evaluate the contribution of recurrent genomic deletions to autosomal recessive conditions, we first mapped all possible loci that are susceptible to recurrent deletions caused by NAHR between directly oriented SDs (Sharp et al. 2006; Dittwald et al. 2013) using the GRCh38 human reference genome sequence (Figure S1, Table S1). The collapsed NAHR map contains 717 unique recurrent deletion regions. We enumerated the subset of recurrent deletion events with available data from screening efforts in the literature or clinical testing to substantiate a prevalence estimate, and focused the subsequent analyses on these genomic intervals (n=51).

We identified 30 deletions with a maximal population prevalence over 1/1,000,000 based upon estimates from the UK Biobank, the Icelandic, and the gnomAD SV studies (Stefansson et al. 2014; Crawford et al. 2019; Collins et al. 2020) (Table 1, Table S2). Of note, these deletion allele frequencies reflect empirical prevalence measurements from adult populations, which closely represent the effective allele frequencies (i.e., combined consideration of both the *de novo* mutation rate and fitness of the variant on a cellular, developmental, and organismal level) suited for recessive disease trait load estimation. These 30 deletions span 64 Mb of unique genomic sequences in the assayable portion of the human genome, contribute to an aggregate population allele burden of 1.3%, and encompass 1517 genes, of which 78 are known to cause recessive disorders. An additional 21 deletions, with populational prevalence possibly lower than 1/1,000,000, are also identified to recur in high prevalence if a clinical cohort is ascertained (Table S2). With the 21 ultra-rare deletions included, the span of genomic coverage increases to 82 Mb; the number of genes involved becomes 1828, with 100 representing established recessive disease trait genes. Moreover, the ‘haploid genetics’ concept begins to emerge as an approach based on observational data and data analyses.

**Table 1.** Recurrent genomic deletions that are prevalent in the population.

| Region | Coordinates (GRCh38) | Population allele frequency (x 10<sup>-6</sup>) | Allele frequency in diagnostic testing (x 10<sup>-6</sup>) | Known recessive genes | Number of coding genes |
| --- | --- | --- | --- | --- | --- |
| 2q13 *NPHP1* | chr2:109930242-110228182 | 5873 | 2616 | *NPHP1* | 3 |
| 15q11.2 | chr15:21311962-23261294 | 2764 | 2287 | - | 14 |
| 16p12.1 | chr16:21754781-22502804 | 592.3 | 627.8 | *OTOA, UQCRC2* | 11 |
| 17p12 HNPP | chr17:14170711-15567588 | 314.8 | 388.6 | *COX10, PMP22* | 9 |
| 16p13.11 | chr16:14772948-16330433 | 312.6 | 433.5 | *NDE1, MYH11, ABCC6* | 15 |
| 1q21.1 BP3-BP4 | chr1:146380249-148811725 | 267.2 | 672.6 | - | 15 |
| 16p11.2 proximal | chr16:29416551-30202090 | 259.6 | 1674 | *PRRT2, ALDOA, TBX6, CORO1A* | 35 |
| 13q12.12 | chr13:22911590-24323812 | 211.7 | 104.6 | *SGCG, SACS, MIPEP* | 7 |
| 22q11.2 LCRA-D | chr22:18530098-21214537 | 177.1 | 4499 | *PRODH, SLC25A1, CDC45, GP1BB, TXNRD2, TANGO2, SCARF2, PI4KA, SNAP29, LZTR1* | 49 |
| 1q21 TAR | chr1:144904297-146209950 | 173.9 | 269.0 | *PEX11B, RBM8A, POLR3GL, HJV* | 22 |
| 10q11.21q11.23 | chr10:45765081-49954967 | 141.2 | 74.73 | *RBP3, ERCC6, SLC18A3, CHAT* | 38 |
| 16p11.2 distal | chr16:28706949-29049993 | 136.1 | 254.1 | *TUFM, ATP2A1, CD19, LAT* | 11 |
| 2q13 | chr2:110494056-112385043 | 128.6 | 149.5 | *ANAPC1, MERTK* | 11 |
| 2q21.1 | chr2:130623447-131386379 | 100.8 | 119.6 | - | 9 |
| 15q13.3 BP4-BP5 | chr15:30246847-32496522 | 93.26 | 896.8 | *FAN1, TRPM1, OTUD7A* | 13 |
| 2q11.2 | chr2:95759114-97430329 | 73.10 | 74.73 | *ADRA2B, NCAPH, LMAN2L, CNNM4* | 24 |
| 17q12 | chr17:36300613-38034442 | 68.86 | 463.4 | *ZNHIT3, PIGW* | 21 |
| 7q11.23 | chr7:73089294-74862006 | 39.35 | 1375 | *NCF1* | 27 |
| 15q11q13 BP3-BP4 | chr15:28580349-30417865 | 35.29 | 134.5 | *NSMCE3* | 10 |
| 3q29 | chr3:195963652-197626678 | 22.69 | 194.3 | *TFRC, PCYT1A, TCTEX1D2, RNF168, NRROS, CEP19* | 23 |
| 17q11.2 | chr17:30621877-32037969 | 22.66 | 149.5 | - | 14 |
| 22q11.2 LCRD-H | chr22:21206521-24255497 | 12.60 | 59.79 | *IGLL1* | 45 |
| 8p23.1 | chr8:7596999-12344083 | 10.08 | 134.5 | *RP1L1, FDFT1* | 50 |
| 10q23 | chr10:79733715-87254783 | 7.562 | 59.79 | *MAT1A, CDHR1* | 31 |
| 17p11.2 Smith Magenis Syndrome | chr17:16777950-20450859 | 5.041 | 687.6 | *TNFRSF13B, ATPAF2, MYO15A, MEIF2, TOP3A, GRAP, B9D1, ALDH3A2* | 48 |
| 15q11 Prader-Willi/ | chr15:21976318- | 2.521 | 687.6 | *OCA2, HERC2* | 26 |
| Angelman syndromes BP1-BP3 | 28537425 |  |  |  |  |
| 15q24 BPA-BPC | chr15:72628218-75278711 | 2.521 | 74.73 | *BBS4, STRA6, EDC3, MPI, COX5A* | 40 |
| 15q11q13 BP3-BP5 | chr15:28569118-32447357 | 2.521 | 59.79 | *NSMCE3, FAN1, TRPM1* | 21 |
| 7q11.23 distal | chr7:75456184-76629927 | 2.521 | 29.89 | *POR, MDH2* | 19 |

Regions are listed in descending order by population prevalence. Genes in the “Known recessive gene” column are ordered by coordinate map positions. Even though it is the third highest NAHR-mediated deletions, the Xp22.31-*STS* deletion is not included in this table because the current list focuses on autosomal recessive conditions. The gene *OTUD7A* in 15q13.3 BP4-BP5 is not reported as a recessive disease at the time of this study; however, patient analysis results from this study support this gene as a candidate recessive disease.

We then catalogued, based on existing knowledge and datasets, a compendium of all reported and predicted carrier alleles for each known recessive trait gene in the human genome. Our objective for this recessive allele catalog is to estimate and dissect the impact of the new mutation recurrent genomic deletions’ contribution to the overall disease burden. Based on mode of inheritance curations from OMIM, DECIPHER, and ClinGen (data accessed on 1/4/2021), a totality of 2657 recessive disease trait genes were assembled. The carrier allele burden for each ‘recessive trait gene’ was calculated by summing up frequencies of unique alleles for all high-quality pathogenic variants from ClinVar, all structural variants (SV) predicted to be LoF from gnomAD SV v2.1, all high-confidence LoF small variants identified in gnomAD v3.1, and the NAHR-mediated recurrent genomic deletions, if applicable. An aggregate of 85,063 small variant and large deletion carrier alleles were identified for the 2657 genes (Table S3). For the 78 known recessive genes in the NAHR deletion regions, the number of per gene pathogenic alleles range from 1 to 308, with a median of 14. As a comparison, the remaining 2510 known rare recessive disease trait genes have a similar median per gene pathogenic allele count, 13, but a wider range, from 1 to 3562.

A limitation of this calculation is that SNV pathogenic missense, in-frame indel, or intronic variants not currently reported in ClinVar are inadvertently omitted. However, we argue that carrier alleles not represented in ClinVar tend to have lower allele frequencies and thus do not have a major impact on the subsequent carrier burden estimates. We further argue that the alleles that receive an entry and curation in ClinVar have higher frequencies – and therefore greater impact on recessive disease, and these are the alleles more easily ascertained in screening tests of clinical diagnostic laboratories. This latter contention is supported by the aggregate gene-level carrier allele burden from our analysis matching empirical experience in genetic testing carrier screenings results (Table S4).(Haque et al. 2016)

Nevertheless, to account for potential unrepresented alleles from recessive disease trait genes that have not been scrutinized by large-scale systematic clinical or research screening, in the subsequent analyses, we supplemented the disease allele pool for each gene with a 10% extra variant load, comprised of ten hypothetical variants each accounting for 1% of the overall carrier burden (see Methods) for each gene. Of note, the NAHR deletion alleles rank as the most (47/78) or second most (12/78) frequent (highest population allele frequency) carrier alleles together comprising over three quarters of known recessive trait genes within NAHR regions! Even with the above-mentioned conservative ‘padding’ to represent ten hypothetical alleles not yet ascertained, the NAHR alleles still contribute to greater than 10% of the total gene-level carrier allele burden for 58 of the 78 genes (Table S5).

### NAHR-deletions contribute a major fraction of recessive disease load to genes mapping within rearrangement hotspots

It is important to note that, for a recessive trait, the population frequency and relativized frequency of a particular allele from a pool of alleles (**Fa**, fraction of allele burden) is not linearly correlated with the probability of sampling a patient with the specific allele from a group of patients (**Fd**, fraction of the disease burden). The distribution of alleles in affected individuals is determined by the pairwise allele frequency products in a pool.

Thus, we calculated allelic contributions to recessive disease load using an *n* x *n* Punnett square, where *n* is the number of carrier alleles for a recessive disease trait gene. The calculated NAHR-deletion contribution to disease can be calculated from the matrix. We denote Fd as the modeled probability of sampling individual carrying at least one recurrent deletion allele from a pool of patients affected with the recessive condition caused by the same gene. We empirically considered a gene to be under significant NAHR-deletion burden for population prevalence of the associated recessive disease trait, if the recurrent genomic deletion is expected in greater than 20% of all patients with this recessive disorder. By this definition, 71% (56/78) of NAHR-region recessive genes, which account for 2.11% of all known recessive genes, are under significant NAHR-deletion burden for recessive disease trait prevalence (Table 2)! In the context of the other alleles from the same gene, the disease contribution of the NAHR-deletion (Fd) ranks at the top for 47 genes, and at second place for 12 more genes. The Fd scores of the top-3 alleles are listed in Table 2 and Table S5 to illustrate a snapshot of the allelic architecture for each recessive trait gene.

**Table 2.**
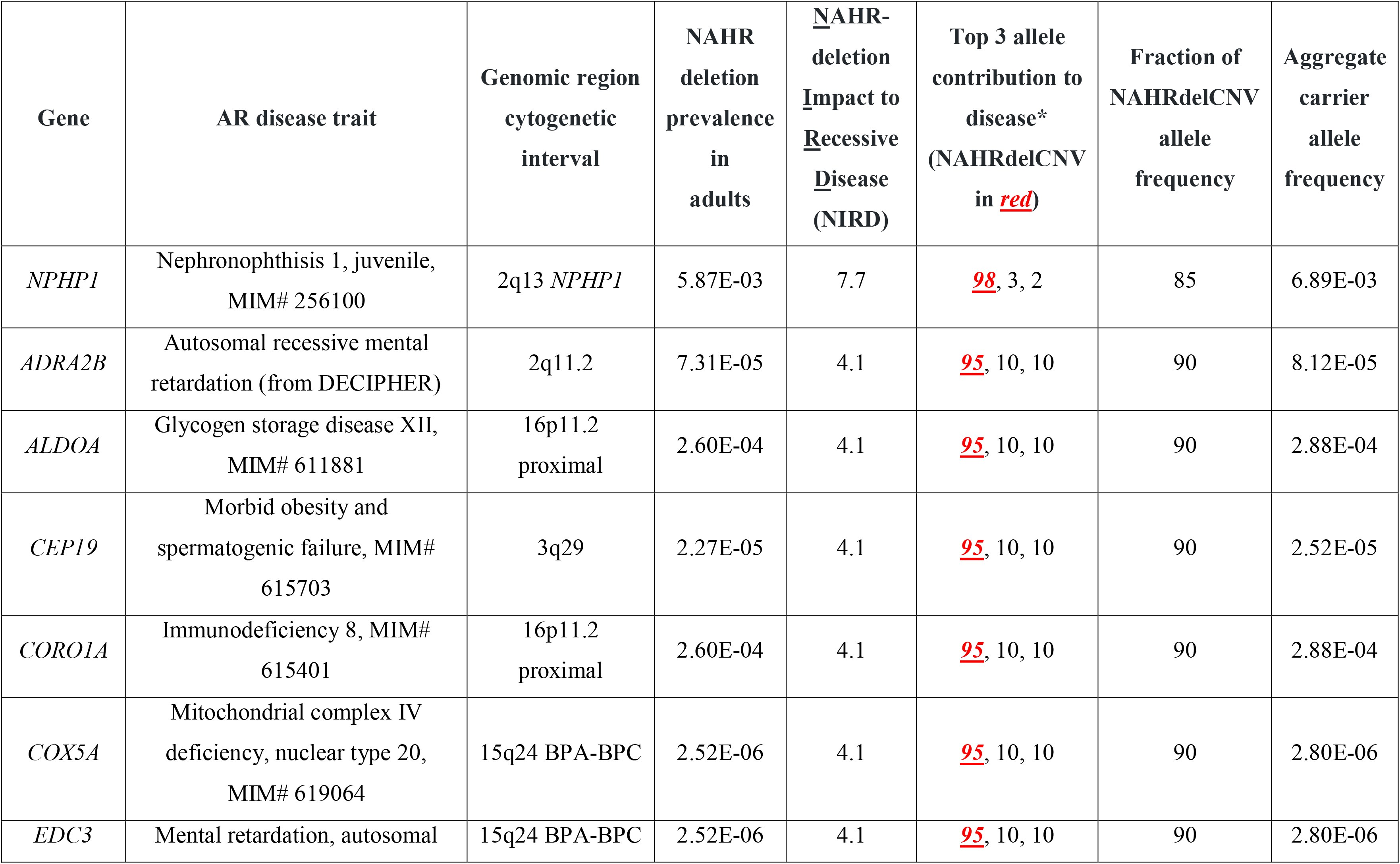
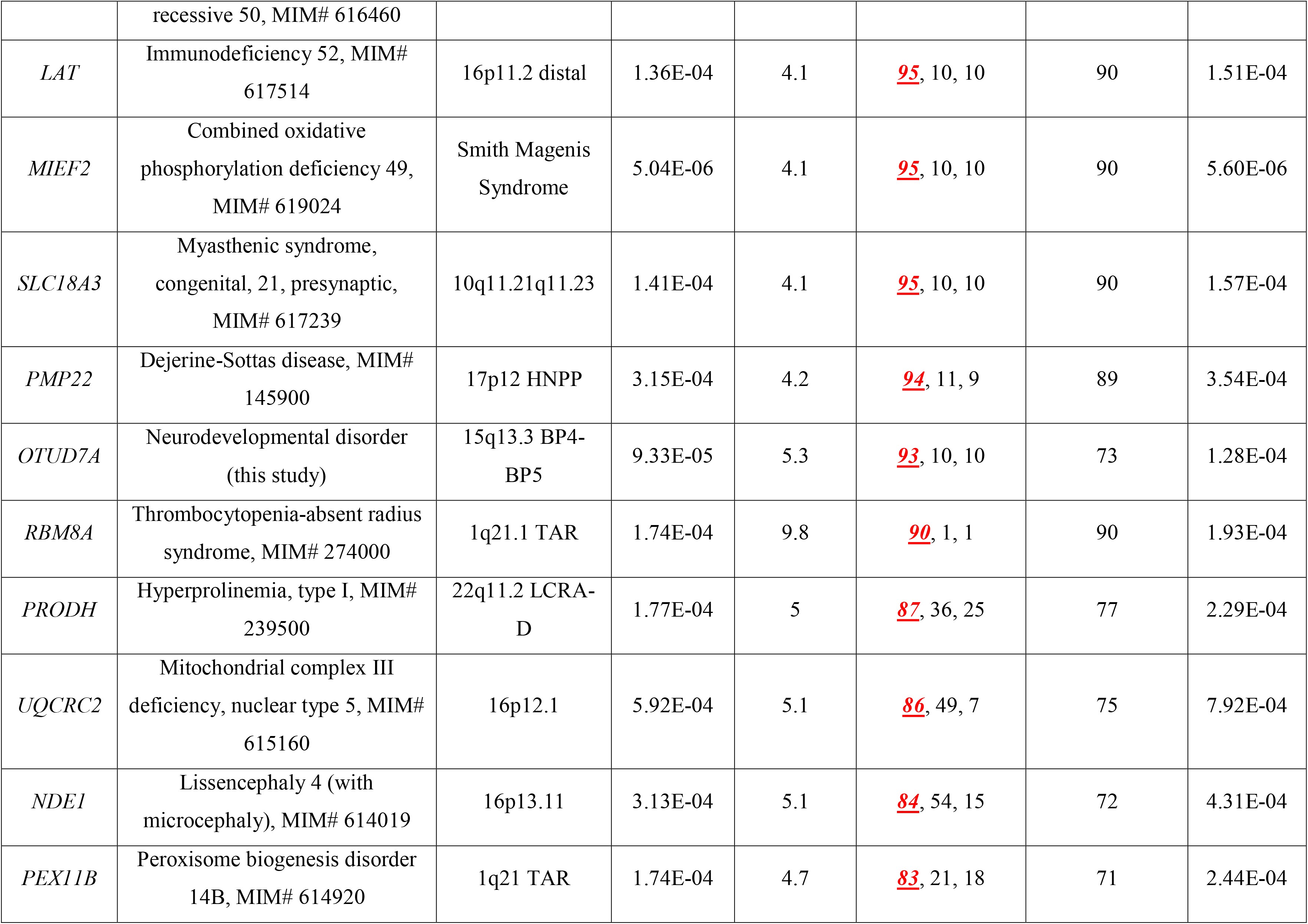
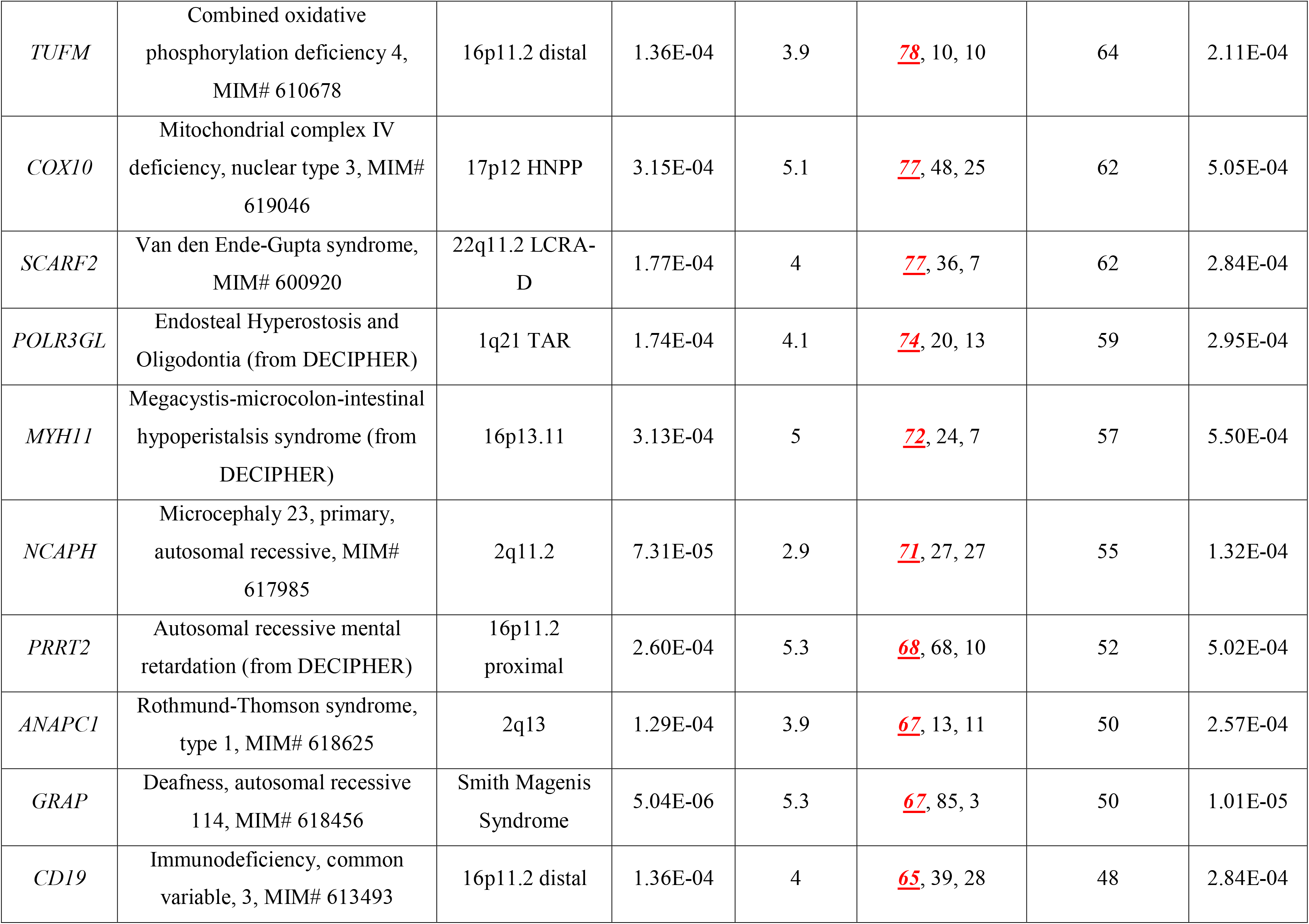
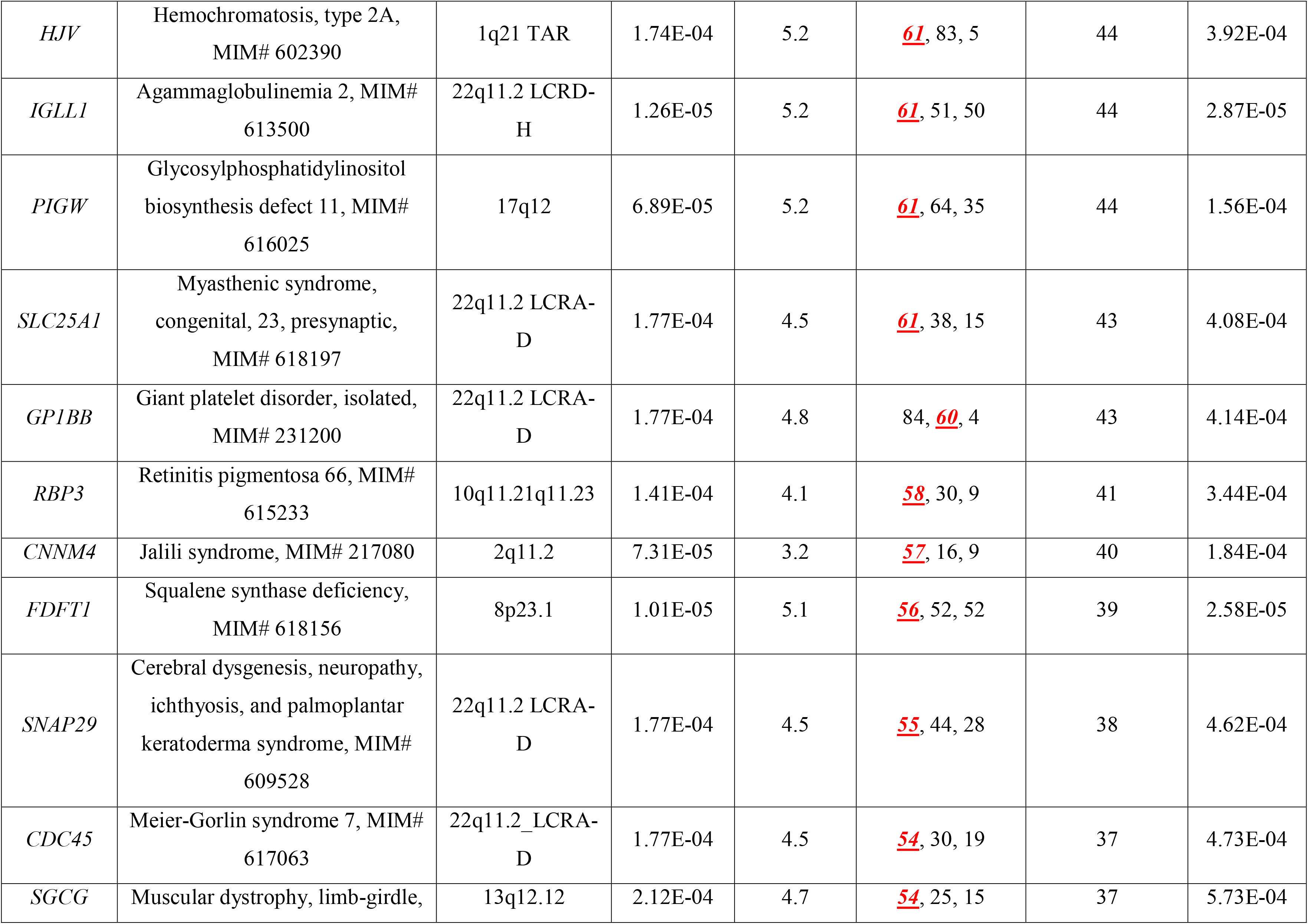
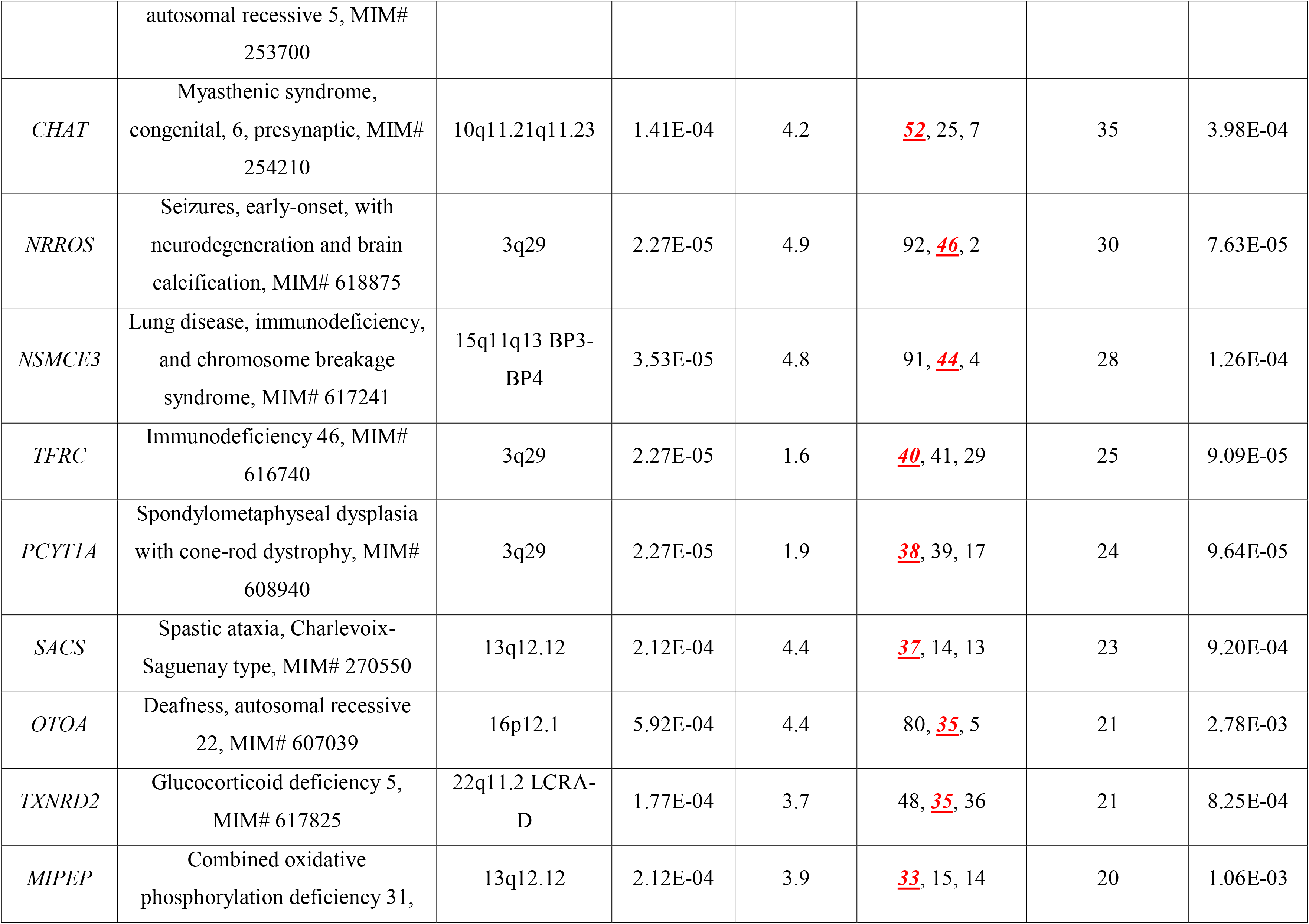
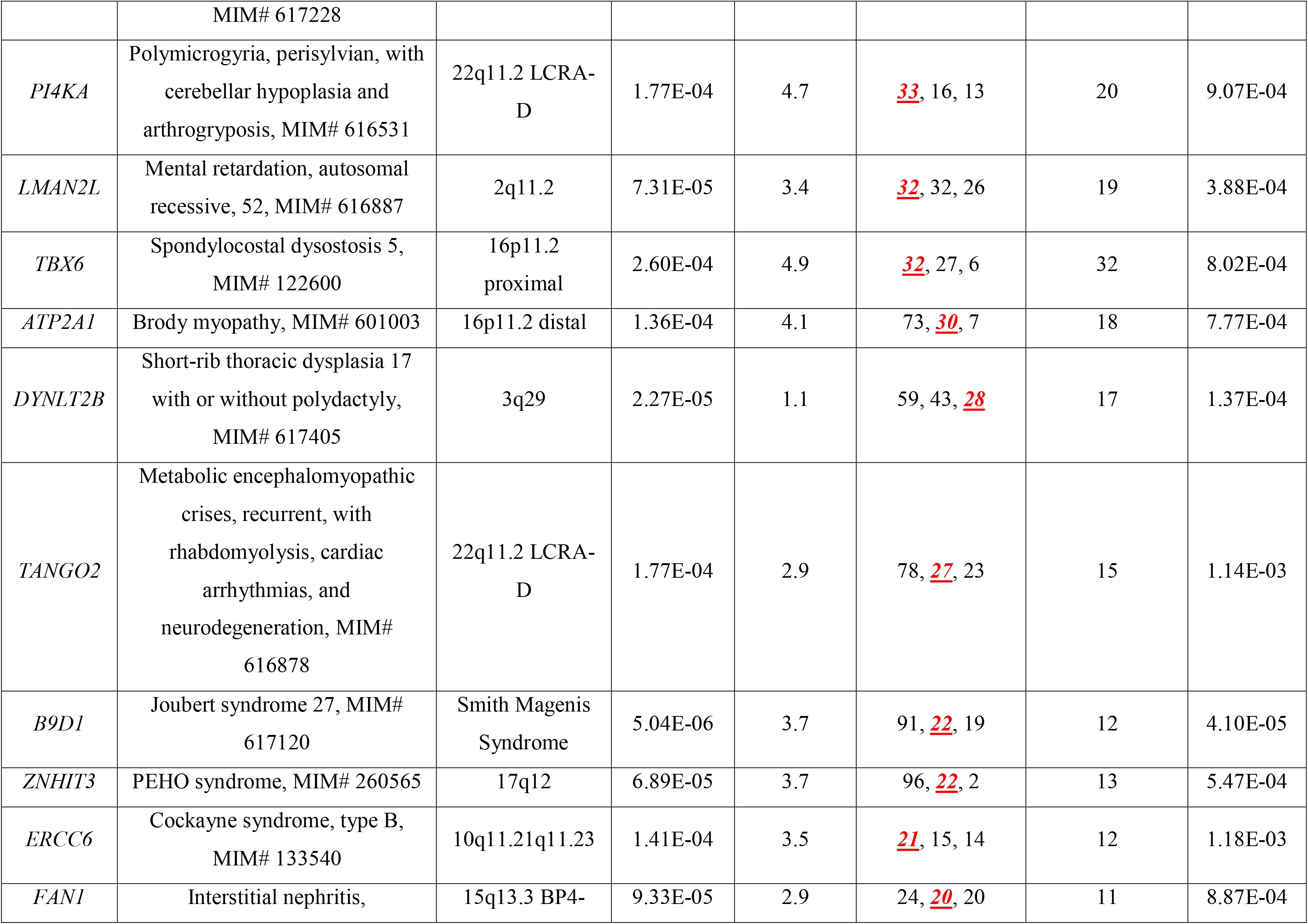
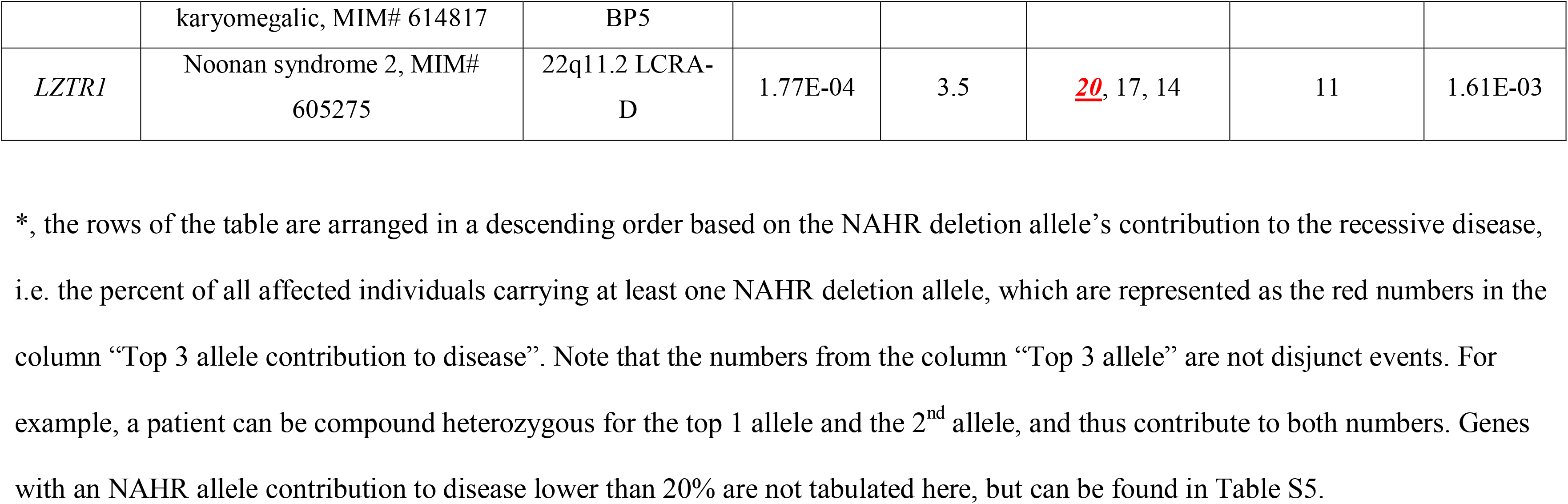
Recessive genes with NAHR-mediated recurrent genomic deletions contributing to more than 20% of the overall disease burden.

| Gene | AR disease trait | Genomic region<br>cytogenetic<br>interval | NAHR<br>deletion<br>prevalence<br>in<br>adults | <u>NAHR</u>-<br>deletion<br>Impact to<br><u>R</u>ecessive<br><u>D</u>isease<br>(NIRD) | Top 3 allele<br>contribution to<br>disease*<br>(NAHRdelCNV<br>in *red*) | Fraction of<br>NAHRdelCNV<br>allele<br>frequency | Aggregate<br>carrier<br>allele<br>frequency |
| --- | --- | --- | --- | --- | --- | --- | --- |
| *NPHP1* | Nephronophthisis 1, juvenile,<br>MIM# 256100 | 2q13 *NPHP1* | 5.87E-03 | 7.7 | *98*, 3, 2 | 85 | 6.89E-03 |
| *ADRA2B* | Autosomal recessive mental<br>retardation (from DECIPHER) | 2q11.2 | 7.31E-05 | 4.1 | *95*, 10, 10 | 90 | 8.12E-05 |
| *ALDOA* | Glycogen storage disease XII,<br>MIM# 611881 | 16p11.2<br>proximal | 2.60E-04 | 4.1 | *95*, 10, 10 | 90 | 2.88E-04 |
| *CEP19* | Morbid obesity and<br>spermatogenic failure, MIM#<br>615703 | 3q29 | 2.27E-05 | 4.1 | *95*, 10, 10 | 90 | 2.52E-05 |
| *CORO1A* | Immunodeficiency 8, MIM#<br>615401 | 16p11.2<br>proximal | 2.60E-04 | 4.1 | *95*, 10, 10 | 90 | 2.88E-04 |
| *COX5A* | Mitochondrial complex IV<br>deficiency, nuclear type 20,<br>MIM# 619064 | 15q24 BPA-BPC | 2.52E-06 | 4.1 | *95*, 10, 10 | 90 | 2.80E-06 |
| *EDC3* | Mental retardation, autosomal recessive 50, MIM# 616460 | 15q24 BPA-BPC | 2.52E-06 | 4.1 | *95*, 10, 10 | 90 | 2.80E-06 |
| *LAT* | Immunodeficiency 52, MIM# 617514 | 16p11.2 distal | 1.36E-04 | 4.1 | <u>95</u>, 10, 10 | 90 | 1.51E-04 |
| *MIEF2* | Combined oxidative phosphorylation deficiency 49, MIM# 619024 | Smith Magenis Syndrome | 5.04E-06 | 4.1 | <u>95</u>, 10, 10 | 90 | 5.60E-06 |
| *SLC18A3* | Myasthenic syndrome, congenital, 21, presynaptic, MIM# 617239 | 10q11.21q11.23 | 1.41E-04 | 4.1 | <u>95</u>, 10, 10 | 90 | 1.57E-04 |
| *PMP22* | Dejerine-Sottas disease, MIM# 145900 | 17p12 HNPP | 3.15E-04 | 4.2 | <u>94</u>, 11, 9 | 89 | 3.54E-04 |
| *OTUD7A* | Neurodevelopmental disorder (this study) | 15q13.3 BP4-BP5 | 9.33E-05 | 5.3 | <u>93</u>, 10, 10 | 73 | 1.28E-04 |
| *RBM8A* | Thrombocytopenia-absent radius syndrome, MIM# 274000 | 1q21.1 TAR | 1.74E-04 | 9.8 | <u>90</u>, 1, 1 | 90 | 1.93E-04 |
| *PRODH* | Hyperprolinemia, type I, MIM# 239500 | 22q11.2 LCRA-D | 1.77E-04 | 5 | <u>87</u>, 36, 25 | 77 | 2.29E-04 |
| *UQCRC2* | Mitochondrial complex III deficiency, nuclear type 5, MIM# 615160 | 16p12.1 | 5.92E-04 | 5.1 | <u>86</u>, 49, 7 | 75 | 7.92E-04 |
| *NDE1* | Lissencephaly 4 (with microcephaly), MIM# 614019 | 16p13.11 | 3.13E-04 | 5.1 | <u>84</u>, 54, 15 | 72 | 4.31E-04 |
| *PEX11B* | Peroxisome biogenesis disorder 14B, MIM# 614920 | 1q21 TAR | 1.74E-04 | 4.7 | <u>83</u>, 21, 18 | 71 | 2.44E-04 |
| *TUFM* | Combined oxidative phosphorylation deficiency 4, MIM# 610678 | 16p11.2 distal | 1.36E-04 | 3.9 | 78, 10, 10 | 64 | 2.11E-04 |
| *COX10* | Mitochondrial complex IV deficiency, nuclear type 3, MIM# 619046 | 17p12 HNPP | 3.15E-04 | 5.1 | 77, 48, 25 | 62 | 5.05E-04 |
| *SCARF2* | Van den Ende-Gupta syndrome, MIM# 600920 | 22q11.2 LCRA-D | 1.77E-04 | 4 | 77, 36, 7 | 62 | 2.84E-04 |
| *POLR3GL* | Endosteal Hyperostosis and Oligodontia (from DECIPHER) | 1q21 TAR | 1.74E-04 | 4.1 | 74, 20, 13 | 59 | 2.95E-04 |
| *MYH11* | Megacystis-microcolon-intestinal hypoperistalsis syndrome (from DECIPHER) | 16p13.11 | 3.13E-04 | 5 | 72, 24, 7 | 57 | 5.50E-04 |
| *NCAPH* | Microcephaly 23, primary, autosomal recessive, MIM# 617985 | 2q11.2 | 7.31E-05 | 2.9 | 71, 27, 27 | 55 | 1.32E-04 |
| *PRRT2* | Autosomal recessive mental retardation (from DECIPHER) | 16p11.2 proximal | 2.60E-04 | 5.3 | 68, 68, 10 | 52 | 5.02E-04 |
| *ANAPC1* | Rothmund-Thomson syndrome, type 1, MIM# 618625 | 2q13 | 1.29E-04 | 3.9 | 67, 13, 11 | 50 | 2.57E-04 |
| *GRAP* | Deafness, autosomal recessive 114, MIM# 618456 | Smith Magenis Syndrome | 5.04E-06 | 5.3 | 67, 85, 3 | 50 | 1.01E-05 |
| *CD19* | Immunodeficiency, common variable, 3, MIM# 613493 | 16p11.2 distal | 1.36E-04 | 4 | 65, 39, 28 | 48 | 2.84E-04 |
| *HJV* | Hemochromatosis, type 2A,<br>MIM# 602390 | 1q21 TAR | 1.74E-04 | 5.2 | <u>61</u>, 83, 5 | 44 | 3.92E-04 |
| *IGLL1* | Agammaglobulinemia 2, MIM#<br>613500 | 22q11.2 LCRD-H | 1.26E-05 | 5.2 | <u>61</u>, 51, 50 | 44 | 2.87E-05 |
| *PIGW* | Glycosylphosphatidylinositol<br>biosynthesis defect 11, MIM#<br>616025 | 17q12 | 6.89E-05 | 5.2 | <u>61</u>, 64, 35 | 44 | 1.56E-04 |
| *SLC25A1* | Myasthenic syndrome,<br>congenital, 23, presynaptic,<br>MIM# 618197 | 22q11.2 LCRA-D | 1.77E-04 | 4.5 | <u>61</u>, 38, 15 | 43 | 4.08E-04 |
| *GP1BB* | Giant platelet disorder, isolated,<br>MIM# 231200 | 22q11.2 LCRA-D | 1.77E-04 | 4.8 | 84, <u>60</u>, 4 | 43 | 4.14E-04 |
| *RBP3* | Retinitis pigmentosa 66, MIM#<br>615233 | 10q11.21q11.23 | 1.41E-04 | 4.1 | <u>58</u>, 30, 9 | 41 | 3.44E-04 |
| *CNNM4* | Jalili syndrome, MIM# 217080 | 2q11.2 | 7.31E-05 | 3.2 | <u>57</u>, 16, 9 | 40 | 1.84E-04 |
| *FDFT1* | Squalene synthase deficiency,<br>MIM# 618156 | 8p23.1 | 1.01E-05 | 5.1 | <u>56</u>, 52, 52 | 39 | 2.58E-05 |
| *SNAP29* | Cerebral dysgenesis, neuropathy,<br>ichthyosis, and palmoplantar<br>keratoderma syndrome, MIM#<br>609528 | 22q11.2 LCRA-D | 1.77E-04 | 4.5 | <u>55</u>, 44, 28 | 38 | 4.62E-04 |
| *CDC45* | Meier-Gorlin syndrome 7, MIM#<br>617063 | 22q11.2_LCRA-D | 1.77E-04 | 4.5 | <u>54</u>, 30, 19 | 37 | 4.73E-04 |
| *SGCG* | Muscular dystrophy, limb-girdle, autosomal recessive 5, MIM#<br>253700 | 13q12.12 | 2.12E-04 | 4.7 | <u>54</u>, 25, 15 | 37 | 5.73E-04 |
| *CHAT* | Myasthenic syndrome,<br>congenital, 6, presynaptic, MIM#<br>254210 | 10q11.21q11.23 | 1.41E-04 | 4.2 | 52, 25, 7 | 35 | 3.98E-04 |
| *NRROS* | Seizures, early-onset, with<br>neurodegeneration and brain<br>calcification, MIM# 618875 | 3q29 | 2.27E-05 | 4.9 | 92, 46, 2 | 30 | 7.63E-05 |
| *NSMCE3* | Lung disease, immunodeficiency,<br>and chromosome breakage<br>syndrome, MIM# 617241 | 15q11q13 BP3-BP4 | 3.53E-05 | 4.8 | 91, 44, 4 | 28 | 1.26E-04 |
| *TFRC* | Immunodeficiency 46, MIM#<br>616740 | 3q29 | 2.27E-05 | 1.6 | 40, 41, 29 | 25 | 9.09E-05 |
| *PCYT1A* | Spondylometaphyseal dysplasia<br>with cone-rod dystrophy, MIM#<br>608940 | 3q29 | 2.27E-05 | 1.9 | 38, 39, 17 | 24 | 9.64E-05 |
| *SACS* | Spastic ataxia, Charlevoix-Saguenay type, MIM# 270550 | 13q12.12 | 2.12E-04 | 4.4 | 37, 14, 13 | 23 | 9.20E-04 |
| *OTOA* | Deafness, autosomal recessive<br>22, MIM# 607039 | 16p12.1 | 5.92E-04 | 4.4 | 80, 35, 5 | 21 | 2.78E-03 |
| *TXNRD2* | Glucocorticoid deficiency 5,<br>MIM# 617825 | 22q11.2 LCRA-D | 1.77E-04 | 3.7 | 48, 35, 36 | 21 | 8.25E-04 |
| *MIPEP* | Combined oxidative<br>phosphorylation deficiency 31, | 13q12.12 | 2.12E-04 | 3.9 | 33, 15, 14 | 20 | 1.06E-03 |
|  | MIM# 617228 |  |  |  |  |  |  |
| *PI4KA* | Polymicrogyria, perisylvian, with cerebellar hypoplasia and arthrogryposis, MIM# 616531 | 22q11.2 LCRA-D | 1.77E-04 | 4.7 | <u>33</u>, 16, 13 | 20 | 9.07E-04 |
| *LMAN2L* | Mental retardation, autosomal recessive, 52, MIM# 616887 | 2q11.2 | 7.31E-05 | 3.4 | <u>32</u>, 32, 26 | 19 | 3.88E-04 |
| *TBX6* | Spondylocostal dysostosis 5, MIM# 122600 | 16p11.2 proximal | 2.60E-04 | 4.9 | <u>32</u>, 27, 6 | 32 | 8.02E-04 |
| *ATP2A1* | Brody myopathy, MIM# 601003 | 16p11.2 distal | 1.36E-04 | 4.1 | 73, <u>30</u>, 7 | 18 | 7.77E-04 |
| *DYNLT2B* | Short-rib thoracic dysplasia 17 with or without polydactyly, MIM# 617405 | 3q29 | 2.27E-05 | 1.1 | 59, 43, <u>28</u> | 17 | 1.37E-04 |
| *TANGO2* | Metabolic encephalomyopathic crises, recurrent, with rhabdomyolysis, cardiac arrhythmias, and neurodegeneration, MIM# 616878 | 22q11.2 LCRA-D | 1.77E-04 | 2.9 | 78, <u>27</u>, 23 | 15 | 1.14E-03 |
| *B9D1* | Joubert syndrome 27, MIM# 617120 | Smith Magenis Syndrome | 5.04E-06 | 3.7 | 91, <u>22</u>, 19 | 12 | 4.10E-05 |
| *ZNHIT3* | PEHO syndrome, MIM# 260565 | 17q12 | 6.89E-05 | 3.7 | 96, <u>22</u>, 2 | 13 | 5.47E-04 |
| *ERCC6* | Cockayne syndrome, type B, MIM# 133540 | 10q11.21q11.23 | 1.41E-04 | 3.5 | <u>21</u>, 15, 14 | 12 | 1.18E-03 |
| *FAN1* | Interstitial nephritis, | 15q13.3 BP4- | 9.33E-05 | 2.9 | 24, <u>20</u>, 20 | 11 | 8.87E-04 |
|  | karyomegalic, MIM# 614817 | BP5 |  |  |  |  |  |
| *LZTR1* | Noonan syndrome 2, MIM#<br>605275 | 22q11.2 LCRA-D | 1.77E-04 | 3.5 | <u>20</u>, 17, 14 | 11 | 1.61E-03 |

\*, the rows of the table are arranged in a descending order based on the NAHR deletion allele's contribution to the recessive disease, i.e. the percent of all affected individuals carrying at least one NAHR deletion allele, which are represented as the red numbers in the column "Top 3 allele contribution to disease". Note that the numbers from the column "Top 3 allele" are not disjunct events. For example, a patient can be compound heterozygous for the top 1 allele and the 2<sup>nd</sup> allele, and thus contribute to both numbers. Genes with an NAHR allele contribution to disease lower than 20% are not tabulated here, but can be found in Table S5.

We next defined a log-scaled index we termed the NAHR-deletion’s Impact to Recessive Disease (NIRD), to depict the gene-level disease load contribution of the NAHR allele relative to an allele with a median level of contribution to the same gene among all population carrier alleles (See Methods section). A positive NIRD score predicts that the NAHR-deletion allele plays a predominant (above the typical allele) role among all carrier alleles of the gene in disease contribution, whereas a negative score predicts a minor (below typical) role. Known recessive genes in the recurrent deletion region tend to have high NIRD scores, with 85% (66/78) scoring above 0, and 74% (58/78) scoring above 2. Of note, the two highest NIRD scores are found in *RBM8A* and *NPHP1*, 9.8 and 7.7, respectively. Both are extremely large values considering the NIRD is log-scaled.

To appreciate the properties of the NIRD scores, we adjusted the algorithm to calculate the disease contribution of any given pathogenic allele for a recessive trait gene, as an Allelic Impact to Recessive Disease (AIRD) score. The most common carrier allele observed in cystic fibrosis, NM_000492.3(*CFTR*):c.1521_1523delCTT (p.Phe508delPhe), also known as the ΔF508 allele, has an AIRD score of 7.3; the third most common carrier allele for Niemann-Pick disease type A, NM_000543.5(*SMPD1*):c.996del (p.Phe333fs), has an AIRD score of 2; a well-known founder mutation observed in ∼10% of patients of Ashkenazi Jewish descent with Tay-Sachs disease, NM_000520.6(*HEXA*):c.1421+1G>C, has an AIRD score of −0.15, due to its lower allele frequency of 1.97 x 10^-5^ in the general population according to gnomAD v3.1.

The NIRD and related findings provide the computational framework that supports two consequences. First, for the ∼2% of known human recessive genes genome-wide or 70% of recessive genes in NAHR regions, one of the most effective but under-utilized approaches and strategies for identifying novel disease-causing alleles from human subjects for these genes is to sequence affected individuals carrying the heterozygous recurrent genomic deletion encompassing the gene of interest. Second, there likely exist uncharacterized recessive disease trait genes that may be most effectively identified by sequencing individuals bearing prevalent recurrent genomic deletions -- i.e. any of the remaining 1439 genes within these deletion regions may have yet to be assigned an AR disease trait and could be novel biallelic/recessive disease trait genes.

### Meta-analysis suggests under-representation of the NAHR-deletion allele in currently discovered recessive disease trait allele pools

The striking prediction of the high contribution of NAHR-deletions to relevant recessive disease trait load is seemingly contradictory to our current impression of the recessive allele landscapes. This implication led us to hypothesize that the NAHR-deletion alleles are currently under-represented in disease characterization efforts. To test this latter hypothesis, we analyzed the distributions of a near-complete catalogue of currently discovered disease alleles in 170 patient families affected with one of the recessive traits whose carrier burden are predicted to be almost exclusively from NAHR-mediated large deletions (Fd > 70% from Table 2). The cohorts are assembled by meta-analysis of all literature reports for patients with the corresponding recessive disease trait disorder recorded in HGMD (version 2020.4), with the assumption that most patients, penetrant for the clinical disease entity, with these extremely rare recessive disease trait disorders characterized in research efforts are reported in the literature. *NPHP1,* the top-ranking gene from Table 2, is a well-characterized recessive trait ‘disease gene’, for which many research characterized patients may not result in published literature. Therefore, *NPHP1* is not included in these analyses because the literature-assembled meta-analysis cohort is unlikely to represent the natural disease allele composition (i.e. clinical practice) in the world.

It is expected that all patients with biallelic disease variants fall into three categories (1) HMZ: those affected with homozygous small variants possibly from a close- or distant-consanguineous relationship, (2) SNV+SNV: those affected with compound heterozygous small variants, and (3) NAHR deletion CNV +SNV (NAHRdelCNV+SNV): those affected with a large deletion in *trans* with a small variant allele. We anticipate that category #1-HMZ accounts for a substantial proportion, demonstrating the well-established robustness of autozygosity mapping as a method for allelic and new recessive trait gene discovery (as populational rare alleles can be escalated to much higher clan allele frequency) (Coban-Akdemir et al. 2020). In outbred pedigrees and populations corresponding to categories #2-SNV+SNV and #3-NAHRdelCNV+SNV, our modeling from the NIRD hypothesis is that #3-NAHRdelCNV+SNV should account for a higher fraction. The opposing trend would suggest that our current disease gene/allele discovery efforts are not exploiting the large deletion allele to the fullest extent that a ‘human haploid genetics’ approach might allow.

All the genes except for *RBM8A* show a poor representation for the #3-NAHRdelCNV+SNV configuration, based on the 93 families when excluding the ones affected with *RBM8A* variants from the entire cohort (Table S6). Note that since most of the variants reported in these families are only documented once in affected human subjects, we cannot rule out the possibility that some of these variants are not causative to the clinical presentation, i.e., the variants of interest are not pathogenic determining alleles. Almost 70% (65/93) of these families carry homozygous disease alleles (52 unique alleles). Based on our modeling and the assumption of random mating, patients with homozygous variants are expected to account for a small fraction of the overall cohort, ranging from 0.53% to 5.4% per gene. However, the observed fractions of homozygotes for each gene are 1.9 to 189 (median 47) fold higher than expected. Furthermore, all of the 65 (or 52 unique) homozygous variants are rare, with 42 being ultra-rare (as defined by not observed in gnomAD v3.1). The collective patterns suggest that current efforts investigating these recessive traits tend to ascertain patients from populations with elevated autozygosity or from targeted population groups with their ethnic-specific disease founder alleles.

To avoid potential confounding factors from study designs and patient ascertainment methods, we removed patients with homozygous variants and focused on those with compound heterozygous variant alleles. Our modeling predicts that for these top-ranking genes analyzed, the number of patients with NAHRdelCNV+SNV should be 2.8 to 20 (median 12) fold higher than the number of patients with compound heterozygous small variants (Table S6). The observed counts from many individual genes are too low to support a meaningful conclusion, but in aggregate, we have identified fewer patients with NAHRdelCNV+SNV (13) compared with patients with compound heterozygous small variants (16). The recurrent deletions involved are 16p13.11 (n=4), 17p12-HNPP (n=3), DiGeorge 22q11.2 (n=2), 10q11.21q11.23 (n=2), 1q21.1-TAR (n=1), and proximal 16p11.2 (n=1). The poor representation of deletion bearing patients shows a bias that under-represents category #3-NAHRdelCNV+SNV and deviates from the expectation driven by our analysis using empirical population allele frequencies and the NIRD score.

*RBM8A* is the only gene from our analysis that demonstrated a discovery pattern favoring #3-NAHRdelCNV+SNV, with the majority (95%, 73/77) of patients affected with the *RBM8A*-thrombocytopenia absent radius (TAR; OMIM #274000) syndrome being compound heterozygous for the 1q21.1-TAR deletion and a small variant, whereas no patients were found to carry homozygous *RBM8A* pathogenic variants (Table S6). This finding is consistent with expectations from our computational modeling based on the allelic spectra illustrating an overwhelming fraction of contribution of the 1q21.1 NAHR-deletion at the disease locus (Tables 2 and S6). Moreover, the observed representation of NAHRdelCNV discovery at this locus, in contrast to other loci, is expected because of a unique characteristic of the *RBM8A*-1q21 locus. The disease presentation requires a combination of the rare 1q21 NAHR-deletion null allele and a common (∼1% minor allele frequency) hypomorphic small variant (Albers et al. 2012). However, neither of the two allele types can be found in patients as homozygotes – the NAHR-deletion homozygotes are lethal and the homozygous hypomorphic small variants are not disease-triggering. The unique molecular allele architecture and disease pathogenic mechanism of *RBM8A*, a condition that is clinically uniform and genetically homogeneous, shuts the door of discovery by sequencing of population with high autozygosity, but spontaneously presented the #3-NAHRdelCNV+SNV configurations for research discovery (Klopocki et al. 2007). Similar expectations, empirical modeling and observations for *Tbx6* derived scoliosis, i.e. *TBX6-*associated congenital scoliosis in mice, were found (Wu et al. 2015; Yang et al. 2019).

### A human haploid genetics and genomics approach to recessive trait genes

We retrospectively analyzed two existing clinical cohorts to find data that test our computational prediction of NAHR-deletions conferring a major disease burden to many recessive disease traits. The configurations of the two cohorts are not optimized for discovery, but seem to have provided preliminary evidence in support of our prediction from computational modeling. The first cohort was assembled focusing on the *COX10* gene, defects of which cause mitochondrial complex IV deficiency (OMIM# 220110) inherited as an AR trait.

*COX10* is located within the 17p12 recurrent deletion that is associated with hereditary neuropathy with liability to pressure palsies (HNPP, OMIM# 162500), a mild form of peripheral neuropathy, or a dominant susceptibility locus to neuropathy after traumatic injury, akin to an animal model observed as the Wallerian degeneration slow phenotype modeled in the *Wld* triplication mouse (Coleman et al. 1998). HNPP is due to decreased dosage of the *PMP22* gene via haploinsufficiency and is inherited as a liability to pressure palsies originally described in the Dutch population and pathologically presenting as tomaculous neuropathy (Pellissier et al. 1987; Lupski 2022); it is often only manifested clinically as multifocal neuropathy elicited after sustained trauma to a peripheral nerve that traverses close to the body surface and manifest as an entrapment neuropathy (Tyson et al. 1996) or an operative carpal tunnel syndrome co-segregating through multiple generations (Potocki et al. 1999). *PMP22* maps within the 1.5 Mb HNPP deletion CNV and *COX10* is the only gene in the deletion interval with a known AR disease trait association other than *PMP22*; the latter *PMP22* is associated with both an AD and AR neuropathy traits (Roa et al. 1993; Shy et al. 2006). Based on our calculation, ∼77% of all patients affected with biallelic *COX10* pathogenic alleles in an outbred population carry one HNPP deletion (Table 2).

We retrospectively investigated results from 596 patients suspected with a mitochondrial disorder who were clinically tested for *COX10* coding region sequencing and deletion/duplication CNV analyses. The strength of the patient assertation strategy from this cohort is that patients were referred based on clinical suspicion and therefore the distribution of pathogenic alleles from this cohort is likely free of a ‘molecular diagnosis bias’. A weakness of this cohort configuration is that the selected disease phenotype is of high genetic heterogeneity, which inherently predicts that only a small number of patients will indeed be affected with a *COX10*-related condition. Nevertheless, we found two patients received a possible molecular diagnostic finding in *COX10,* both carrying the HNPP deletion as one allele.

In subject #1, a hemizygous variant resulting in an in-frame small duplication of two amino acids, c.1277_1282dup (p.M426_L427dup) in exon 7 of *COX10*, was identified in *trans* to the HNPP deletion (Figure S2). Subject #2, whose referral indication is COX deficiency, has a rare VUS c.858G>T (p.W286C) in *COX10* in *trans* with the HNPP deletion. In the remaining patients without a definitive molecular diagnosis, two patients were found to have the heterozygous HNPP deletions, but a second hit in *COX10* was not found, although we cannot rule out the possibility of additional findings in intronic or regulatory regions. These findings, though under-powered, are consistent with our prediction that most patients with cytochrome c oxidase deficiency carry one HNPP deletion allele; either *de novo* or inherited. Considering the high frequency of the HNPP susceptibility allele (Berg et al. 2010) with absence of selection and late adult onset disease (DiVincenzo et al. 2014), it is possible that more novel *COX10* disease alleles can be revealed by sequencing individuals with the HNPP deletion and a mitochondrial spectrum of clinical phenotypes, thereby improving our understanding of the biological function of the *COX10* gene.

The second cohort we assembled is based on the criteria that a patient carries one of the NAHR-deletions and that genotype information of the non-deleted allele is available for analyses. Thus, we identified such individuals from a cohort of 11,091 subjects who were referred for clinical exome sequencing (cES) at a diagnostic laboratory due to a differential clinical diagnosis including various suspected genetic disorders. We performed an initial screen for patients carrying one of the genomic deletions from Table 1, which resulted in 161 subjects carrying one recurrent deletion and 3 subjects carrying two. The two most frequently observed types of deletions, the 15q11.2 BP1−BP2 deletion (n=41) and the *NPHP1*-2q13 deletion (n=23), are excluded from downstream analysis. This exclusion is because none of the coding genes from the 15q11.2 BP1−BP2 deletion have been implicated to be associated with a Mendelian disease trait (Jonch et al. 2019), and the critical gene at 2q13, *NPHP1*, has been already extensively studied (Yuan et al. 2015). We also excluded six subjects harboring the X-linked, hemizygous deletion in the Xp22.31 *STS* locus. After excluding these three groups of deletion CNVs, cES data from personal genomes of 95 subjects, collectively harboring 96 incidences or 26 types of recurrent genomic deletions, were available for us to build the second cohort (Table S7).

This second cohort is not optimized for discovery because it is a collection of various different deletions without any enrichment for a targeted phenotype. Additionally, despite a subset of these patients carry one of the disease-associated large deletions that are known before cES, they are still referred for cES analyses; such a property predicts that the disease pathogenesis mechanism found in this cohort tend to be more complex than a typical Mendelian disease cohort. Such individuals may more likely to be represented by a ‘blended phenotype’ (Posey et al. 2017).

Again, in accordance with our expectations, more than a quarter (26/95) of these subjects were found to have probable small variant molecular diagnostic findings independent from the deletion. From the remaining 69 subjects with an apparent undiagnostic ES result, we identified 4 subjects with rare variants in coding regions exposed by the deletion as potential molecular diagnoses (Table 3). The first patient is subject #1 described above with HNPP deletion and a *COX10* small variant allele.

**Table 3.**
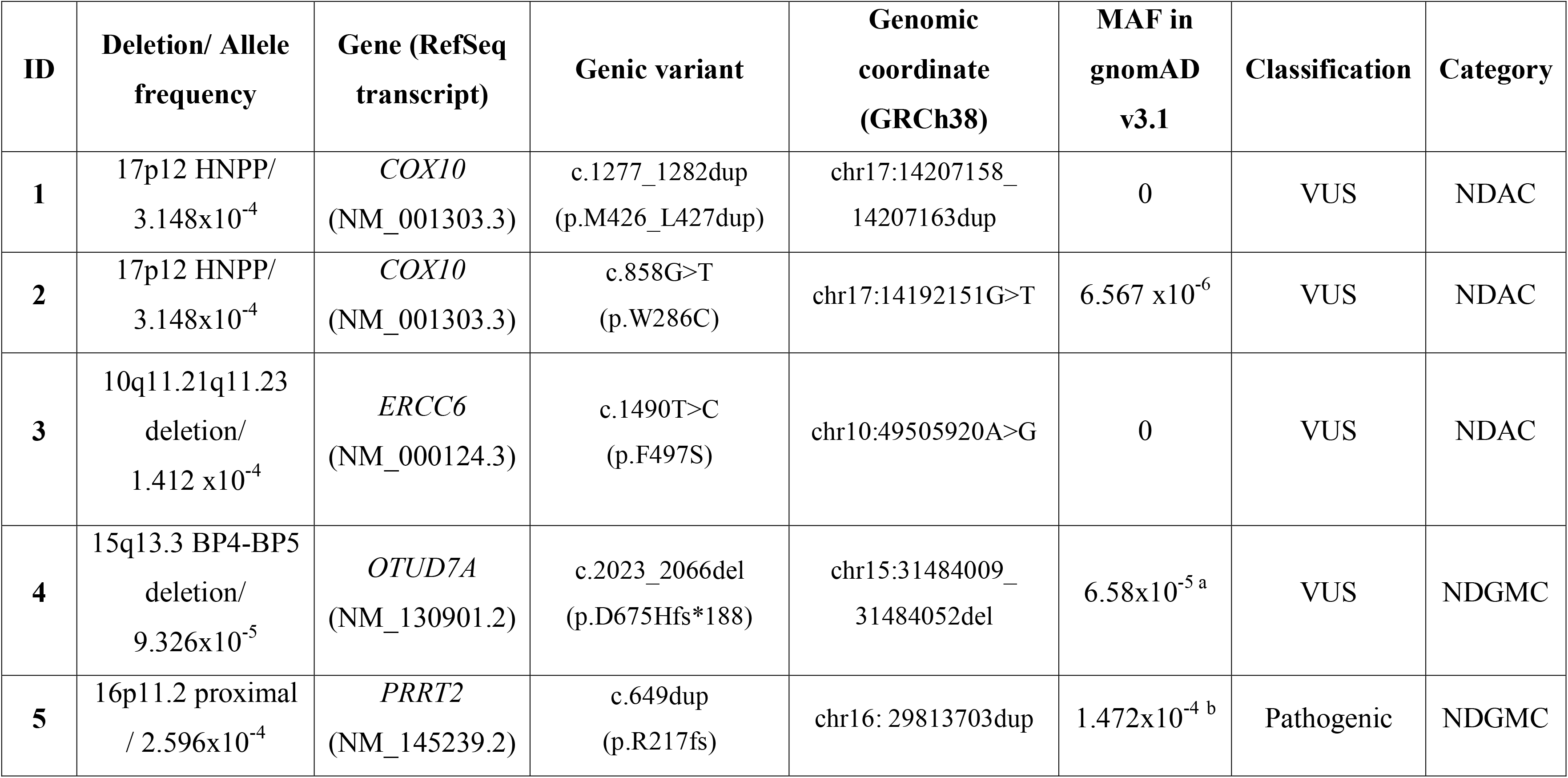
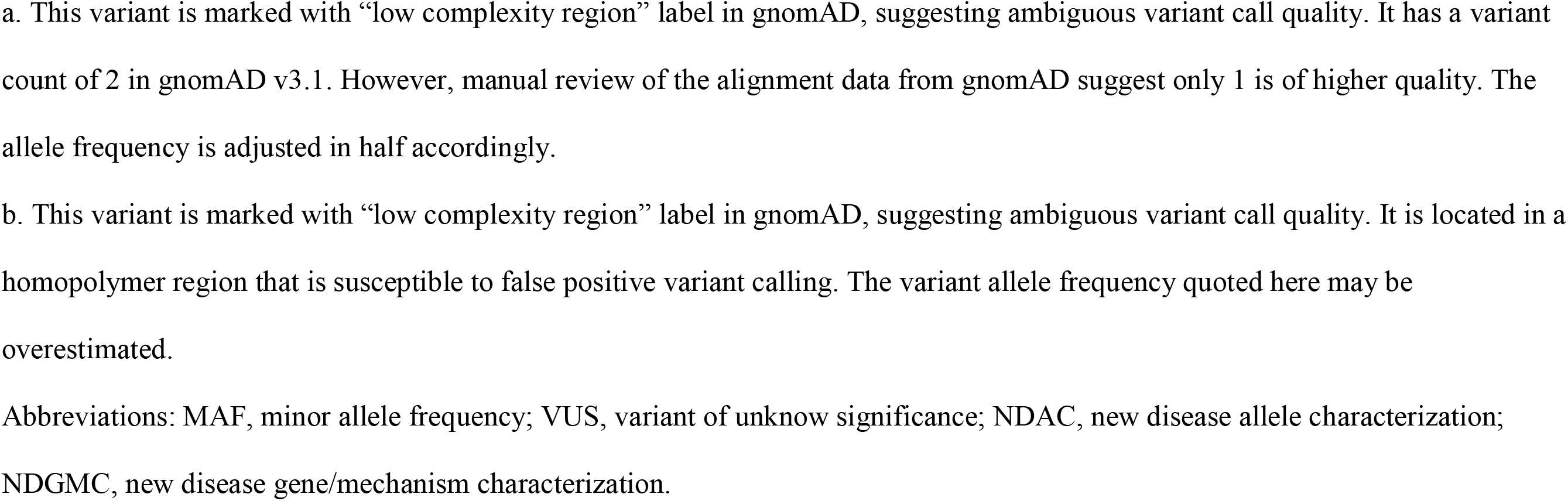
Clinically significant sequence variants uncovered by the deletions. Subjects #1 and #2 were identified in a COX10-phenotype-driven cohort analysis. Subjects #1, #3, and #4 were identified in the molecular-deletion-driven clinical exome data reanalysis.

| **ID** | **Deletion/ Allele frequency** | **Gene (RefSeq transcript)** | **Genic variant** | **Genomic coordinate (GRCh38)** | **MAF in gnomAD v3.1** | **Classification** | **Category** |
| --- | --- | --- | --- | --- | --- | --- | --- |
| **1** | 17p12 HNPP/<br>3.148x10<sup>-4</sup> | *COX10*<br>(NM_001303.3) | c.1277_1282dup<br>(p.M426_L427dup) | chr17:14207158_<br>14207163dup | 0 | VUS | NDAC |
| **2** | 17p12 HNPP/<br>3.148x10<sup>-4</sup> | *COX10*<br>(NM_001303.3) | c.858G>T<br>(p.W286C) | chr17:14192151G>T | 6.567 x10<sup>-6</sup> | VUS | NDAC |
| **3** | 10q11.21q11.23<br>deletion/<br>1.412 x10<sup>-4</sup> | *ERCC6*<br>(NM_000124.3) | c.1490T>C<br>(p.F497S) | chr10:49505920A>G | 0 | VUS | NDAC |
| **4** | 15q13.3 BP4-BP5<br>deletion/<br>9.326x10<sup>-5</sup> | *OTUD7A*<br>(NM_130901.2) | c.2023_2066del<br>(p.D675Hfs*188) | chr15:31484009_<br>31484052del | 6.58x10<sup>-5 a</sup> | VUS | NDGMC |
| **5** | 16p11.2 proximal<br>/ 2.596x10<sup>-4</sup> | *PRRT2*<br>(NM_145239.2) | c.649dup<br>(p.R217fs) | chr16: 29813703dup | 1.472x10<sup>-4 b</sup> | Pathogenic | NDGMC |

- a. This variant is marked with “low complexity region” label in gnomAD, suggesting ambiguous variant call quality. It has a variant count of 2 in gnomAD v3.1. However, manual review of the alignment data from gnomAD suggest only 1 is of higher quality. The allele frequency is adjusted in half accordingly. - b. This variant is marked with “low complexity region” label in gnomAD, suggesting ambiguous variant call quality. It is located in a homopolymer region that is susceptible to false positive variant calling. The variant allele frequency quoted here may be overestimated.
Abbreviations: MAF, minor allele frequency; VUS, variant of unknown significance; NDAC, new disease allele characterization; NDGMC, new disease gene/mechanism characterization.

The second patient, subject #3, has clinical features including ataxia, developmental delay, microcephaly, and short stature. A recurrent 10q11.21q11.23 deletion (Stankiewicz et al. 2012) was identified in *trans* to a novel missense variant allele c.1490T>C (p.F497S) in the *ERCC6* gene. Biallelic variants in *ERCC6* are associated with cerebro-oculo-facio-skeletal syndrome 1 (COFS1, MIM# 214150) or Cockayne syndrome type B (CSB, MIM# 133540). The high allele frequency of the 10q11.21q11.23 deletion (1.412 x10^-4^) increases the probability for a second allele with ultra-low frequency, like the c.1490T>C (p.F497S) *ERCC6* variant, to be correlated with a set of human clinical phenotypes.

Subject #4 presented with severe neurodevelopmental diseases and dysmorphic features. We identified a hemizygous *OTUD7A* frameshift variant allele c.2023_2066del (p.D675Hfs*188) in *trans* with the recurrent 15q13.3 BP4-BP5 deletion, providing evidence for *OTUD7A* as a new disease gene. The recurrent deletion mediated by BP4 and BP5 at the 15q13.3 locus is associated with highly variable NDD (neurodevelopmental disorder) phenotypes, ranging from asymptomatic to mild to moderate intellectual disability, epilepsy, behavioral issues distinct from neurotypical behaviors (e.g. autism spectrum disorders, attention deficit hyperactivity disorders, etc.), and variable dysmorphic features (Ben-Shachar et al. 2009; van Bon et al. 2009). While heterozygous deletion causes highly variable phenotypes, reported homozygous 15q13.3 BP4-BP5 deletion consistently manifest disease phenotypes including significant NDD, epilepsy, hypotonia, visual impairments, and other less common phenotypes including autism spectrum disorder, short stature, failure to thrive, microcephaly and variable dysmorphic features (Table S8) (Endris et al. 2010; Lepichon et al. 2010; Masurel-Paulet et al. 2010; Spielmann et al. 2011; Masurel-Paulet et al. 2014). The critical gene responsible for this ‘ciliopathy like clinical presentation’ of the 15q13.3 BP4-BP5 deletion has been debated, but evidence suggests that *OTUD7A*, encoding a member of a family of deubiquitinating enzymes, may be a plausible candidate (Uddin et al. 2018; Yin et al. 2018).

Studies using syntenic heterozygous deletion mouse models suggest a critical role of Otud7a in neuronal development and brain function (Uddin et al. 2018; Yin et al. 2018). Otud7a-null mouse models manifest many cardinal features of the 15q13.3 deletion syndrome (Yin et al. 2018). The c.2023_2066del (p.D675Hfs*188) variant identified in subject #4 maps to the last exon of the *OTUD7A* gene, and is thus predicted to not result in nonsense-mediated decay (NMD) (Coban-Akdemir et al. 2018). However, the variant is predicted to result in substitution of the C-terminal amino acids after aspartic acid with 187 novel amino acids and a premature termination of the protein translation (PTC). This change may remove the C-terminal Zinc finger A20-type domain and abolish the normal function of the protein. Our finding in Subject #4, together with recent case reports of patients with a homozygous missense *OTUD7A* variant alleles (Garret et al. 2020), or compound heterozygous 15q13.3 deletion in *trans* with a frameshift *OTUD7A* variant (Suzuki et al. 2020), supports our contention and corroborates that *OTUD7A* may be the critical ‘driver gene’ in the 15q13.3 deletion syndrome. *OTUD7A* may be sensitive to gene dosage effect, and contribute to disease etiology at least in part through a biallelic AR disease trait mechanism.

Interestingly, we observe that the population small variant allele pool for *OTUD7A* is depleted for LoF alleles based on gnomAD. Without the 15q13.3 deletion contributing to a major carrier burden, the paucity of small variant disease alleles for *OTUD7A* would make disease association establishment using patient data much more challenging. From an alternative perspective, *OTUD7A*’s current apparent ‘high’ gene intolerance to haploinsufficiency (pLI=0.95) may have incidentally portrayed it as a ‘dominant’ Mendelian disease gene, whereas the calculated high NIRD score (5.3) of the gene strongly indicates that the intolerance to haploinsufficiency should be much lower, i.e. low likelihood of being an AD trait gene.

In Subject #5 with severe NDD, we identified a c.649dup (p.R217fs) pathogenic variant in the *PRRT2* gene in *trans* with the recurrent 16p11.2 BP4-BP5 deletion, providing compelling evidence for a novel disease AR trait inheritance mechanism for *PRRT2*. The 16p11.2 BP4-BP5 recurrent deletion is known to be associated with mild dysmorphisms, macrocephaly, and neuropsychiatric phenotypes including DD/ID and autism spectrum disorder (ASD) with incomplete penetrance, a NDD (Weiss et al. 2008; Shinawi et al. 2010).

The *PRRT2* gene is highly expressed in mouse brain and spinal cord during early embryonic development (Chen et al. 2011). Heterozygous LoF variants in *PRRT2* cause movement and seizure disorders including familial infantile convulsions with paroxysmal choreoathetosis (OMIM# 602066), episodic kinesigenic dyskinesia 1 (EKD1, OMIM# 128200), or benign familial infantile seizures 2 (BFIS2, OMIM# 605751), with incomplete penetrance documented (Meneret et al. 2012). The c.649dup (p.R217fs) allele is the most frequent pathogenic variant, occurring at a mutational hotspot with homopolymer of 9 cytosine bases adjacent to 4 guanine bases that are susceptible to DNA replication errors (Heron et al. 2012). Currently, autosomal dominant (AD) is considered as the only disease inheritance mode for *PRRT2* traits in OMIM, although preliminary evidence from case reports suggest that *PRRT2* can cause a more severe NDD through a biallelic pathogenic mechanism and an AR inheritance model (Labate et al. 2012). Our findings in Subject #5 provide further support the contention of a new rare disease trait type, AR versus AD, and inheritance mechanism due to *PRRT2* biallelic variation. Moreover, these observations may also highlight a potential compound inheritance gene dosage (CIGD) model that explains penetrance of certain neurological phenotypes observed in patients with the 16p11.2 deletion; a similar biallelic compound inheritance gene dosage model underlies the penetrance of ∼10-12% of all congenital scoliosis worldwide (Liu et al. 2019a).

### NAHR-deletions contribute to recessive disease burden in population-specific patterns

As suggested earlier, the contribution of a given allele to rare recessive disease trait burden is influenced by the composition of other pathogenic alleles from the same gene. Although the genetics and genomics fields are beginning to appreciate inter-individual variabilities in NAHR rates associated with alternative genomic structural haplotypes (Shaw-Smith et al. 2006; Wu et al. 2015; Yuan et al. 2015) as well as polymorphisms from *trans* acting factors controlling homologous recombination, such as *PRDM9* (Berg et al. 2010), we currently still assume that NAHR mutation rates at a given locus are relatively constant across different populations and genomic ethnic backgrounds. This potentially leaves the remaining alleles, the small variants, as the major driver for any variability in allelic architecture from different population groups.

To investigate the degree of inter-population variability for small variant recessive alleles, we used ethnic information from gnomAD, and conducted the modeling described earlier for four population groups, African (AFR), Latino (AMR), East Asian (EAS) and European (EUR) (Table S5). Population specific NIRD scores are compared with the general population to generate ΔNIRD (Figure 3), which can be used to inform the relative odds ratio for NAHR-deletions in rare AR disease traits in the specific populations. These analyses provide preliminary computational confirmation for the suspected population variability in NIRD, which implicates that the precision of NIRD can be improved by ‘tuning the disease model’ with population specific allelic architecture. In light of these surprising findings, we tentatively propose, i.e. we hypothesize, that a prioritization strategy based on prior knowledge of population allele frequency spectra can be applied to enhance discovery in research study design of genomic sequencing among individuals with large recurrent deletions. We cautiously note that some of the population groups analyzed here may not have a sufficient sample size to allow a complete representation of disease alleles of relatively lower frequency, which may result in overestimation of ΔNIRD when the score is positive. Additional population specific allele frequency data are warranted to improve the accuracy of these analyses.

**Figure 3.**
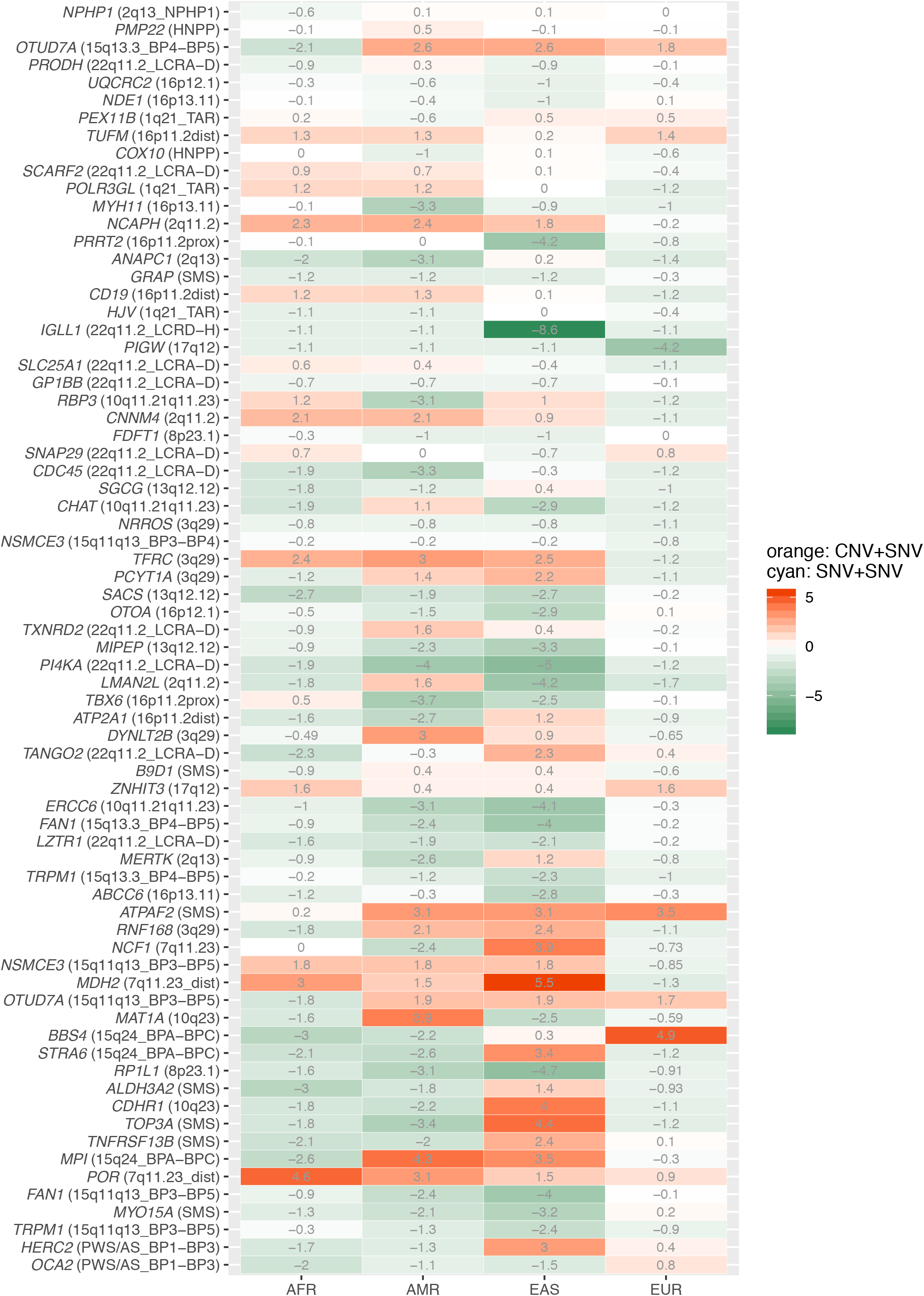
ΔNIRD scores for a population group relative to the general population. ΔNIRD is calculated by subtracting the general population NIRD from the specific population group NIRD. A positive ΔNIRD score suggests an even higher recessive disease contribution of the NAHR-deletion from that population group. A negative ΔNIRD score indicates a relatively higher probability for an affected patient to carry biallelic small variants in that population group. Genes that have zero ΔNIRD scores for all four population groups are not depicted in this heatmap.

## Discussion

We cataloged the population frequency spectra of pathogenic alleles across all known AR disease traits. Our analyses highlighted the high frequency of SV mutagenesis and NAHR deletion alleles in genomic instability regions susceptible to this type of structural variant mutagenesis mediated rearrangement (Lupski 2007). We subsequently computationally interrogated the impact of allele frequency distributions on recessive disease trait burden using a modified Punnett square. We used this computational and genetic approach to consider the impact of the NAHRdelCNV allele on recessive disease traits relative to other recessive trait biallelic variants. Our findings indicate the potentially dramatic disease impact of these NAHRdelCNV – a group of new mutation alleles resulting from recurrent rearrangement; our findings have profound implications for disease biology and molecular diagnosis worldwide.

Considering current clinical genomics practice and research investigations, we postulate that the role of recurrent genomic deletions and new mutation contributing to recessive diseases is under-appreciated, therefore potentially impeding discovery of new disease genes and alleles. Traditionally, researchers and clinicians tend to consider large causative genomic deletions as dominant disease trait halpoinsufficient alleles, with the driver dosage sensitive gene(s) mapping within the deleted interval. This haploinsufficiency assumption likely arose because these alleles were almost all identified as heterozygotes by screening in symptomatic cohorts and frequently as *de novo* CNV mutations. Our analysis of population frequency implicates that the heterozygous state is often not disease causing, and therefore that these variants act as contributors to recessive disease when they occur in *trans* to an AR disease trait allele.

Although many recurrent deletions contribute considerable carrier allele burden to individual recessive Mendelian disease traits, these deletions are often large enough to include other genes whose homozygous depletions are incompatible with live birth. Two exceptions are the 2q13-*NPHP1* deletion and the *SMN1* deletion (the *SMN1* deletion CNV, observed to be found as a carrier state in outbred populations, is predicted by our analyses to be the most frequent NAHR-mediated deletion, NAHRdelCNV allele as shown in Table S1). The fact that other large recurrent deletions are almost never observed in patients as homozygous losses may have led investigators to overlook their equally important role in contributing to recessive disease traits as compound heterozygotes (Boone et al. 2013).

Clinically, there has not been a consensus on whether exome/genome sequencing should be pursued after a recurrent genomic deletion has been identified in a patient. Some may argue that, under the assumption that most Mendelian diseases are caused by one “unifying diagnosis”, the identification of a large genomic deletion can be evidence to ‘demote’ additional candidate molecular diagnoses in the same patient. Our data argue the opposite: that in patients with a recurrent “contiguous gene deletion syndrome”, the possibility of revealing an “additional” recessive disease trait molecular diagnosis cannot be ignored.

Of note, four of the five hemizygous small variants exposed by the deletion in Table 3 are located in repeat or difficult-to-sequence regions. The *COX10* and the *PRRT2* frameshift variants were incorrectly ‘called’ as heterozygous changes by the original exome variant calling pipeline due to challenges of calling indels and potentially other ‘mappability’ issues inherent to such regions of the human genome. If heightened diligence for variant falling in these difficult to assay variation regions were not taken to examine specifically look into the deleted hemizygous interval, these molecular diagnostic variants could have easily been missed during analyses. We suggest this contention might be one of the reasons contributing to the proposed under-detection of recurrent deletion (i.e. NAHRdelCNV) + small variant cases for recessive trait disorders.

Of note, the NIRD score devised in this study not only directly depicts the NAHRdelCNV contribution to the gene’s rare recessive trait disease load, but also highlights features not captured by the haploinsufficiency tolerance score pLI. Of the 48 genes with a high NIRD (>3.5) score, 6 have high pLI scores (>0.9) predicting intolerance to haploinsufficiency. These pLI estimations are not accurate because the calculations failed to factor in the population prevalence of NAHR-deletion alleles. Gene level constraint scores should be adjusted particularly for the high NIRD genes.

Taken together, we suggest that the future of new recessive disease trait genes and allele discovery will greatly benefit from two approaches: (1) genomic sequencing of individuals with recurrent deletions (NAHRdelCNV) and (2) sequencing in population of elevated autozygosity (Figure 4). The autozygosity mapping approach has been the classic approach for new disease gene discovery in medical genetics. This approach enables an adept strategy to target patient populations (by assessing their degree of autozygosity from social and family histories) in order to assemble the appropriate cohorts for research investigations. However, disease trait alleles revealed by this approach are usually limited to a specific population.

**Figure 4.**
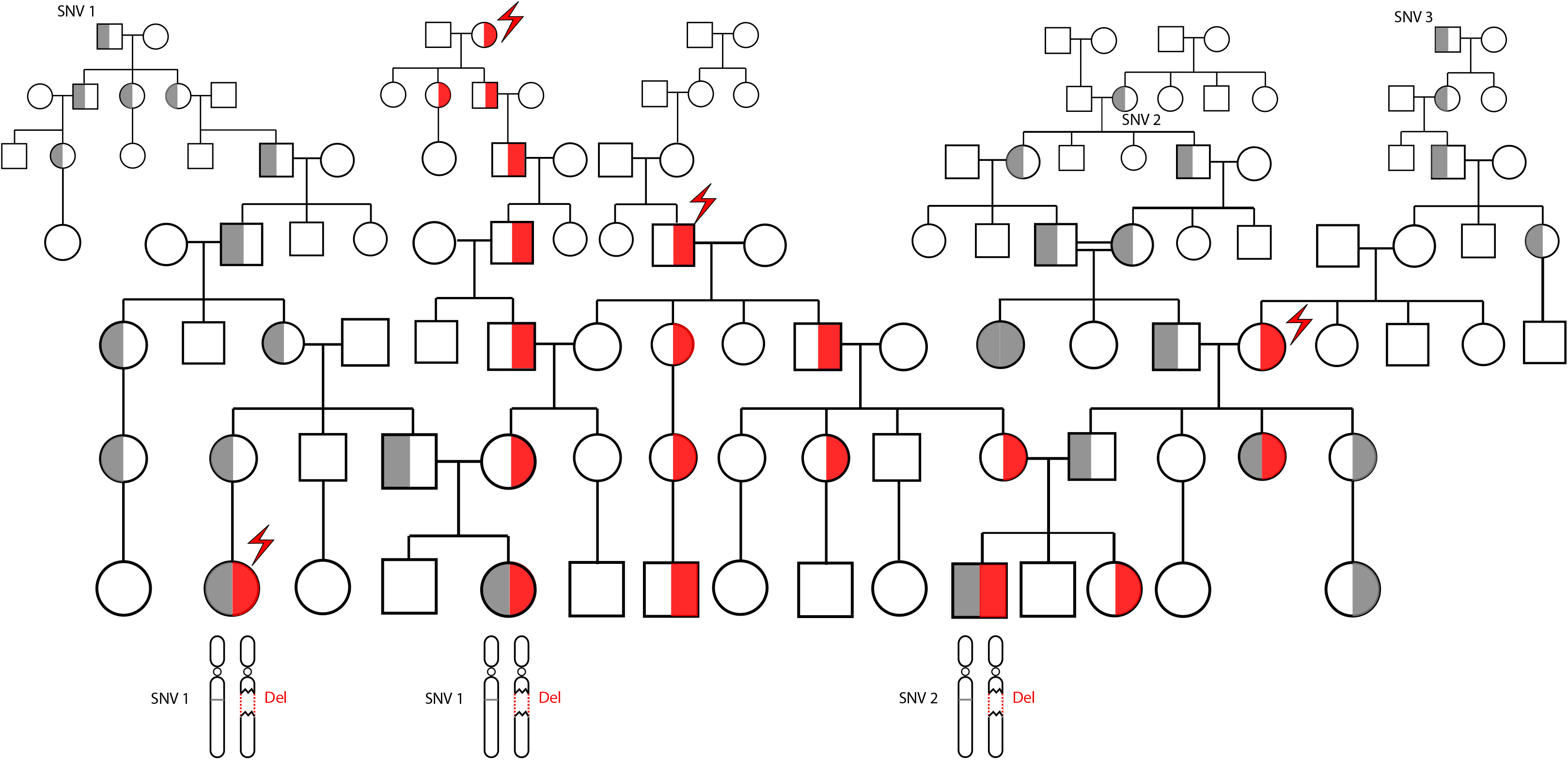
A ‘segmental haploid genomics’ approach for characterization of new disease genes or alleles for autosomal recessive conditions. The illustration above depicts dynamics of disease alleles in an autosomal recessive condition whose collective carrier burden is contributed by a recurrent genomic deletion (red area) and a few single nucleotide variants (SNVs, grey area). Individuals affected with biallelic pathogenic changes frequently carry the deletion as one of the two alleles, because the deletion arises recurrently in multiple lineages (indicated by the red lightning bolt arrow). SNV disease alleles for recessive genes tend to be passed on from ancestral generations, and may drift away without being noticed if they do not converge with another disease allele (SNV3). However, they may emerge to medical attention frequently in families with high degree of autozygosity, as illustrated in generations 3 and 4 for SNV2.

The NAHR-deletion sequencing approach, on the other hand, capitalizes on the known high mutation rates of NAHRdelCNV and has the potential to assign clinical significance to alleles independent of ethnic backgrounds, this latter contention at least partly owing to the recurrent and high mutation rate of the NAHR SV mutagenesis mechanism. Nevertheless, this approach requires prior knowledge and screening of individuals for the recurrent deletion CNVs. It may have been challenging to collect a cohort of patients with large recurrent deletions two decades ago, but the ‘clinical awareness’ of genomic disorders (Lupski 2021; Lupski 2022), advancements in clinical testing and populational screening have made such a genomic experimental effort feasible, either from large diagnostic centers or from clinical registries (Weiss et al. 2008; Shinawi et al. 2010; Mannik et al. 2015; Dewey et al. 2016; Crawford et al. 2019; Jonch et al. 2019; Edwards et al. 2021). Even when available subject numbers are limited for recurrent genomic deletion CNVs with extremely low penetrance, it is possible to tune the disease gene/allele characterization strategy by targeting specific phenotypes, as demonstrated at the Smith Magenis Syndrome -*MYO15* locus two decades ago (Liburd et al. 2001).

Whilst researchers are starting to sequence cohorts of individuals with large ‘Mb-sized’ deletions (Egloff et al. 2018; Beck et al. 2019; Zhao et al. 2020) and performing SNV and CNV analysis in one combined WGS assay, it is imperative for clinical and diagnostic genomicists to foster guidelines that facilitate routine genomic sequencing (ES or WGS) on patients who are found to have recurrent genomic deletions, i.e. NAHRdelCNV, that will benefit both the patients and the research human subjects worldwide. Given the great potential in the near future for disease gene discoveries within intervals of genomic disorder deletion CNVs, these patients will benefit from routine or perhaps even more prioritized reanalysis of sequencing data (Liu et al. 2019b).

Performing DNA sequencing on personal genomes with higher population prevalence of recurrent deletions (those from Table 1) carries additional long-term promise for clinical characterization of common variant alleles, extending the current scope of focus on mono- or bi-allelic inheritance into the more complex spectrum of disease inheritance – including the compound inheritance gene dosage model, CIGD (Wu et al. 2015). High prevalence recurrent genomic deletions are often associated with a high degree of incomplete penetrance of disease phenotype, ranging from 10-90% (Rosenfeld et al. 2013). The disease causal mechanism described in this study, for example, the 16p11.2 deletion + the *PRRT2* small variant leading to NDD, may explain a small portion of the previously-attributed missing heritability for the disease “penetrance” at 16p11.2. However, the totality of the missing heritability is likely not explained by the recessive trait model alone, because the observed disease penetrance is likely to exceed the aggregation of the recessive disease allele prevalence (individually rare) on the non-deleted chromosome. It is plausible that alternative disease models exist, in which the critical gene triggers disease presentation when one rare LoF allele is combined with one or a set of milder hypomorphic alleles with common population frequency. This compound inheritance model has been demonstrated at the *RBM8A*-1q21.1 locus in association with the TAR syndrome (Albers et al. 2012), the *TBX6*-16p11.2 locus in association with congenital scoliosis (Wu et al. 2015; Yang et al. 2019), the *F12*-5q35 Sotos deletion locus in association with blood clotting (Kurotaki et al. 2005), and the *TBX4-FGF10* lung disease (Karolak et al. 2019).

Our data and analyses in this study was focused on coding sequence changes. Moreover, the limited size of patient cohort ascertained for each recurrent deletion CNV may decrease the power of identifying high frequency hypomorphic alleles. These will be dramatically empowered by genome sequencing in a larger patient cohort. Nevertheless, the unifying theme for both the strictly recessive model, and the more complex compound inheritance gene dosage model, is that large recurrent, NAHR derived, genomic deletion CNVs, i.e. NAHRdelCNV alleles often associated with a genomic disorder when heterozygous (Lupski 2022), may provide a unique perspective in characterization of new disease trait loci and alleles, the biology of disease, and the emerging field of human haploid genetics.

## Methods

### Construction of a genome-wide map for NAHR-mediated recurrent genomic deletions

Possible loci for recurrent deletions were identified by enumerating all regions flanked directly oriented low copy repeat (LCR) pairs, LCR sometimes referred to as segmental duplication (SD) in the human genome. These LCR pairs stimulate NAHR-derived deletions of the genomic intervals mapping between the directly oriented pairs. A genomic interval containing the same set of genes can be flanked by different LCR pairs. LCR pairs clustering to the same NAHR region were computationally identified and reduced to generate the coordinates of the merged NAHR intervals.

Metrics that could inform estimation of the new mutation rates for each genomic disorder were kept for each SD elements from the merged cluster of repeats, including repeat lengths, distance in between, and sequence similarity (Liu et al. 2011). Gene content of the deleted segment is expected to influence the fitness of this allele. We used the number of genes with a high pLI score, i.e. greater intolerant to haploinsufficiency, to estimate the level of selection to the genomic deletion in the population. These metrics were used to calculate a score to estimate the relative populational prevalence of these CNVs. Coordinates for deletion breakpoints are calculated as a weighted average of all the ranges of possible SDs that may mediate the deletion. Thus, the coordinates are not precise predictions for a specific deletion observed in individual patients, but rather average of all possible types of deletions that were collapsed into the merged deletion. Two separate NAHR-deletion maps were generated using SegDup tracks from GRCh38 and GRCh37.

To account for the issue that relying on a reference human genome haplotype may lead to underrepresentation of recurrent deletion, we performed comprehensive literature review to search for recurrent deletions that only occur on alternative haplotypes. We found that the chromosome 17q21.31 recurrent deletion, which is known to occur on an alternative inversion haplotype (Koolen et al. 2006; Lupski 2006), is only represented in an alternative contig, chr17_GL000258v2_alt. Thus, this genomic region was manually patched to the analysis result as detailed in the GitHub code. We cannot exclude that additional recurrent deletion regions similar to the 17q21.31 deletion may be missed from this analysis if they are not well represented in the literature.

### Prevalence curation for NAHR-mediated recurrent genomic deletions

For genomic deletions with a population prevalence over 1/1,000,000, prevalence estimates were calculated based on the UK Biobank cohort (Crawford et al. 2019) and the Icelandic cohort (Stefansson et al. 2014). If the prevalence estimate was not significantly different between these cohorts (Fisher’s exact test, *p*>0.05), the estimate based on the UK Biobank data was taken.

Otherwise, prevalence from a third cohort [gnomAD SV (Collins et al. 2020) or other region specific literature] was compared with estimates from the UK Biobank and the Icelandic cohorts, and the group with a closer match was taken.

For genomic deletions with a prevalence lower than 1/1,000,000, we investigated a cohort of 33,452 patients who were referred for clinical Chromosomal Microarray Analysis (CMA) using custom designed Agilent oligo-based Comparative Genomic Hybridization arrays (Yuan et al. 2020). Of note, deletion prevalence estimates from the CMA cohort do not represent actual prevalences in the general population, but can inform relative prevalence comparison among rare variant mutational events in the population.

### Recessive disease carrier burden calculation

High-quality ClinVar variants were defined as having a pathogenic or likely pathogenic label with at least one-star review status (accessed 01/21/2021). LoF variants from gnomAD SV were defined by variants meeting all the following criteria (1) PASS filter in the VCF file with a quality score over 500, (2) PROTEIN_CODING LOF flag or PROTEIN_CODING DUP_LOF flag in the VCF file, (3) POPMAX allele frequency lower than 1% and no homozygote counts, (4) less than 80% of the span overlapping with segmental duplications, and (6) the LoF consequence affects all RefSeq transcripts of a recessive gene.

High-confidence LoF small variants from gnomAD v3.1 were defined by variants that fulfill all the following criteria: (1) PASS filter from the gnomAD v3.1 VCF file, (2) do not fall into a low complexity region, (3) QUALapprox score lower than 1×10^5^, (4) sequenced in over 7.5×10^4^ alleles, (5) population allele frequency lower than 1% with no homozygous counts, and (6) marked as a high-quality LoF variant by LOFTEE (Karczewski et al. 2020).

### Calculation of NAHR-deletion contribution to disease burden for a specific recessive disorder

All the following calculations are based on the concept of random mating by sampling from a pool of alleles. Suppose that at an autosomal recessive trait locus, we have *n* alleles, A_1_, A_2_, …, A_k_, …, A_n_, with allele frequencies *p_1_*, *p_2_*, …, *p_k_*, …, *p_n_*. Without loss of generality, we nominate the k^th^ allele as the NAHR-deletion allele -our allele of interest - and let the others index the small variant alleles. For most large recurrent deletion CNVs, homozygous loss of the deletion is incompatible with live birth.

The two exceptions are the 2q13-*NPHP1* deletion and the 15q13.3 BP4-BP5 deletion, for which homozygous deletions are compatible with live birth. Also, the 15q13.3 BP4-BP5 deletion is encompassed by the 15q11q13 BP3-BP5 deletion, so the enclosed recessive genes have two NAHRdelCNV alleles contributing to them.

Hypomorphic variant alleles that cause disease when they co-occur in combination with LoF alleles, which we denote as A_h_, are not disease-causing in the homozygous states. Such alleles are observed in *RBM8A* and *TBX6*, but at present not in others. The exceptions described above regarding *RBM8A*, *TBX6*, the 2q13-*NPHP1* deletion, and the 15q13.3 BP4-BP5 deletion have been accounted for in the calculations for these special circumstances. For simplicity, they are not incorporated in the equations below for illustration, although the modifications to account for them are simple adjustments to the sums and formulas presented below. The modified equations used in the modeling are described in the supplemental methods as well as reflected in the online code.

We denote the probability of an individual carrying the NAHR-deletion allele is

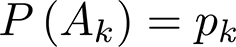

The contribution of the NAHR-deletion to the allele load (**Fa**) is the fraction of the NAHR-deletion allele frequency over the sum of all allele frequencies across all functional alleles -except for the hypomophric alleles that do not in themselves cause disease.

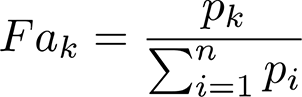

The Punnett square below models the expected frequencies of an individual to be affected with the recessive trait given the population frequencies of each pair of alleles under a random mating model.

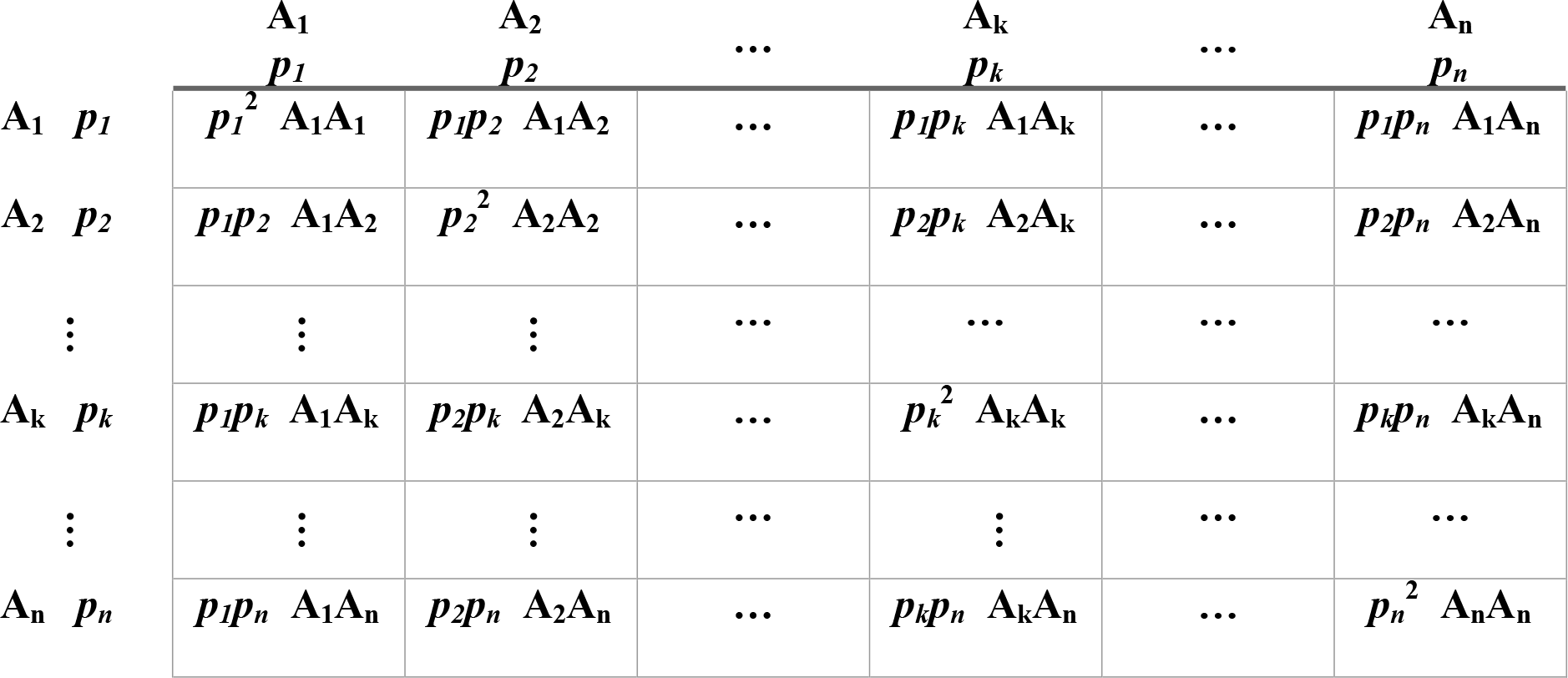

| | $A_1$<br>$p_1$ | $A_2$<br>$p_2$ | ... | $A_k$<br>$p_k$ | ... | $A_n$<br>$p_n$ |
| --- | --- | --- | --- | --- | --- | --- |
| $A_1$ $p_1$ | $p_1^2 A_1 A_1$ | $p_1 p_2 A_1 A_2$ | ... | $p_1 p_k A_1 A_k$ | ... | $p_1 p_n A_1 A_n$ |
| $A_2$ $p_2$ | $p_1 p_2 A_1 A_2$ | $p_2^2 A_2 A_2$ | ... | $p_2 p_k A_2 A_k$ | ... | $p_2 p_n A_2 A_n$ |
| ⋮ | ⋮ | ⋮ | ... | ... | ... | ... |
| $A_k$ $p_k$ | $p_1 p_k A_1 A_k$ | $p_2 p_k A_2 A_k$ | ... | $p_k^2 A_k A_k$ | ... | $p_k p_n A_k A_n$ |
| ⋮ | ⋮ | ⋮ | ... | ⋮ | ... | ... |
| $A_n$ $p_n$ | $p_1 p_n A_1 A_n$ | $p_2 p_n A_2 A_n$ | ... | $p_k p_n A_k A_n$ | ... | $p_n^2 A_n A_n$ |

The probability for an individual to be affected with the recessive disorder is the sum of pairwise products of all carrier alleles with contribution from the homozygous NAHR-deletion lethal allele subtracted

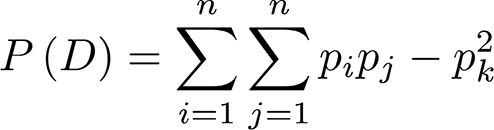

The probability for an individual to be both affected with the recessive disorder and carrying the NAHR-deletion is

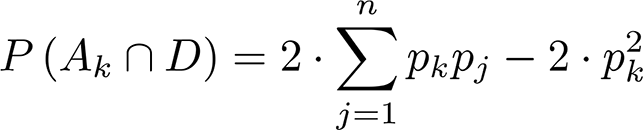

The contribution of individuals with the NAHR-deletion to the recessive disease load (**Fd**) is

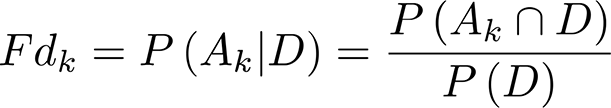

The odds for an individual with the recessive disease to carry the NAHR deletion is

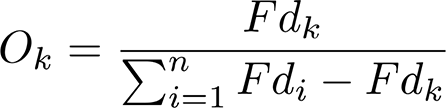

Note that the sum of Fd across alleles *1*, *2*, *3*, …, *n* equals the sum of the Punnett square matrix plus the lower triangular and the upper triangular. This is equivalent to summing up homozygous allele products once plus compound heterozygous allele products twice. This characteristic arises because the events of recessive disease involving the k^th^ and the j^th^ allele are not disjoint and they overlap for the compound heterozygote entries in the Punnett square.

To calculate the NAHR-deletion Impact to Recessive Disease score, the odds of the NAHR allele is compared to that of the “median” allele from the same gene.

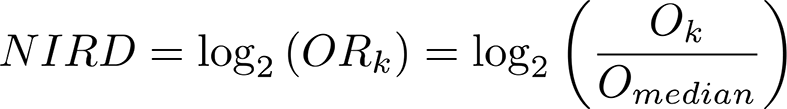

The “median” allele is defined as the midpoint of remaining alleles in the same gene that comprise a cumulative sum of the top 90% of the overall sum of Fd. The alleles consisting the lower 10% of overall Fd sum are disregarded because (1) we find many genes have a long tail of ultra-rare alleles without a frequency estimate from gnomAD, and (2) we supplemented 10% of hypothetical alleles to each gene in our analysis.

### Curation of recessive disease trait alleles from the meta-analysis of the 170 patients identified from the literature

The Human Gene Mutation Database (HGMD, version 2020.4) was queried for recessive disease trait alleles in genes for which ≥70% of the disease carrier burden was attributed to NAHR-mediated large deletions; i.e. NAHRdelCNV. The literature linked to each disease allele that was marked as a disease-causing mutation (DM) or possible DM (DM?) was mined for the allelic state of the variant (homozygous, compound heterozygous with another SNV, or compound heterozygous with the NAHR-mediated deletion, NAHRdelCNV) as well as the number of unrelated families carrying the allele. Homozygous variants were counted once per family, while compound heterozygous SNVs received one count each per family. Gross deletions and insertions larger than 50 bp were excluded. *RBM8A* was the only gene for which disease-associated polymorphisms, with additional supporting functional evidence (DFP), were also included; DM/DM? alleles as compound heterozygous with DFP variants were dismissed for all genes other than *RBM8A*. *MYH11* and *PMP22* are the only two genes in this curation effort to have disease associations of both autosomal dominant (AD) trait and autosomal recessive trait inheritance. *MYH11* alleles were only added if they were associated with hypoperistalsis. Similarly, only biallelic loss-of-function *PMP22* alleles were included. The total population frequency of each curated allele was retrieved using gnomAD v3.1.

### Retrospective analyses of extant cES for a novel rare recessive disease trait gene and allele discovery from individuals with recurrent genomic deletions

This study has been performed in accordance with the research protocol approved by Institutional Review Boards at Baylor College of Medicine. A waiver of informed consent has been obtained (H-41191). The patients were evaluated by clinical exome sequencing (cES), with sequencing, data analysis, and interpretation procedures described previously (Yang et al. 2013; Yang et al. 2014). The identification of deletion CNVs was based on a SNP array platform, which was performed concurrently with the exome assay (Dharmadhikari et al. 2019). Sanger dideoxy DNA sequencing was performed as a validation method for candidate diagnostic small variant alleles.

### Code availability

Detailed code for generation of the NAHR-deletion map, preparation of disease trait alleles, and calculation of disease burden is available at https://github.com/liu-lab/cnvNAHR/.

## Data Availability

Detailed code for generation of the NAHR-deletion map, preparation of disease alleles, and calculation of disease burden is available at https://github.com/liu-lab/cnvNAHR/.

## Acknowledgments

This work was supported in part by the National Human Genome Research Institute (NHGRI) grant number R35HG011311 to PL, Baylor College of Medicine Precision Medicine Initiative Pilot Award to PL, National Human Genome Research Institute (NHGRI)/ National Heart Lung and Blood Institute (NHLBI) grant number UM1HG006542 to the Baylor Hopkins Center for Mendelian Genomics (BHCMG), and the National Institute of Neurological Disorders and Stroke (NINDS) R35NS105078 to JRL.

## Supplemental Figure and Table legends

**Figure S1. Genome-wide map for all predicted NAHR recurrent genomic deletions.** Each predicted deletion event is marked as a green horizontal bar below the chromosome ideograms. The vertical bars above the chromosome ideograms illustrates the density for segmental duplications in a 1000-bp moving window.

**Figure S2.** Compound heterozygous HNPP deletion and *COX10* variant leading to recessive COX10 deficiency in Subjects #2 and #3. **A.** The *COX10* gene spans the repeat sequence that mediate the recurrent HNPP deletion at chromosome 17p12. The *COX10* variant in the Subject #2 is located at the 3’ end of the *COX10* gene on exon 7, which is inside the HNPP deletion interval. The *COX10* variant in the Subject #3 is located at the *COX10* gene exon 7, which is embedded in a CMT1A-REP. Red segments, exons of the *COX10* gene; yellow arrows, CMT1A-REPs; thunderbolts, *COX10* variants observed in Subject #2 and #3, respectively. **B.** Diagram illustrating the scheme of the relationship between the SNV/indel and recurrent deletion identified in Subject #2 and #3.

**Table S1. All 717 recurrent genomic deletions predicted based on the repeat structure in the human reference genome GRCh38.**

**Table S2. All clinically reported recurrent deletions and their prevalance estimates.**

**Table S3. Carrier disease allele frequencies by allele.**

**Table S4. Carrier allele frequency burden by gene.** The list is ranked by genes from the highest burden to the lowest burden. The frequency burden in this list only includes the actual observed variants; the 10% extra hypothetical uncharacterized alleles as described in the Methods section are not included.

**Table S5. Gene-level NAHR contribution to carrier allele and recessive disease burden as well as NAHR Deletion Impact to Recessive Traits (DIRT).** This table is comprised of five panels, representing results generated using data from the general population and four specific ethnic groups, including African (AFR), Latino (AMR), East Asian (EAS), and European (EUR).

**Table S6. Meta-analyses for literature reported patients affected with the 13 recessive disorders contributed by significant NAHR-mediated deletion burden.** Compound heterozygous variants are split into two rows with each row representing one variant.

**Table S7. Molecular findings of recurrent deletions identified from clinical exome sequencing.**

**Table S8. Literature review for 15q13.3 recurrent deletions.**

