## Supplementary figures and images for "Sequencing individual genomes with recurrent genomic disorder deletions: an approach to characterize genes for autosomal recessive rare disease traits"

### Supplemental Figure 1

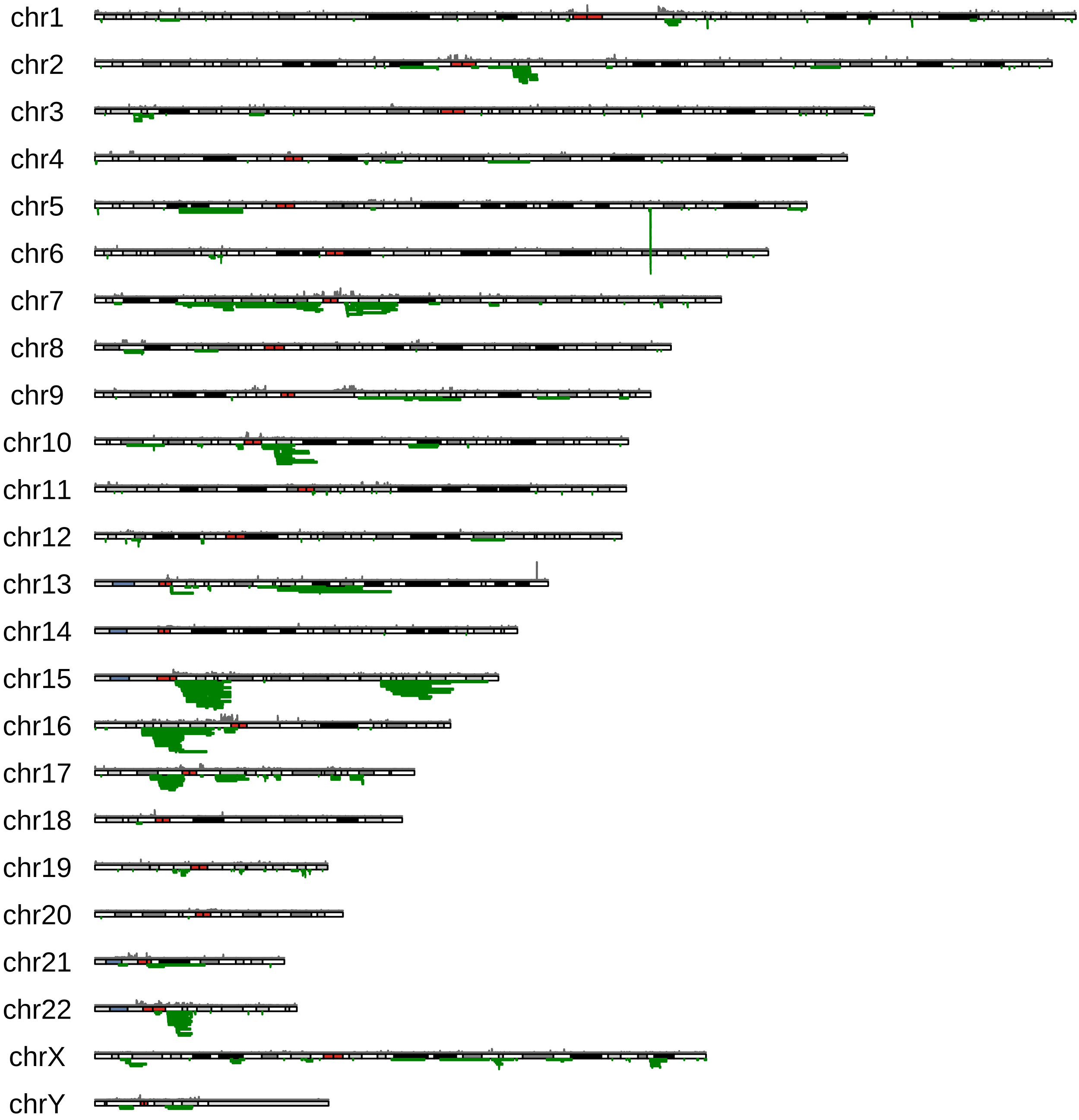

### Supplemental Figure 2

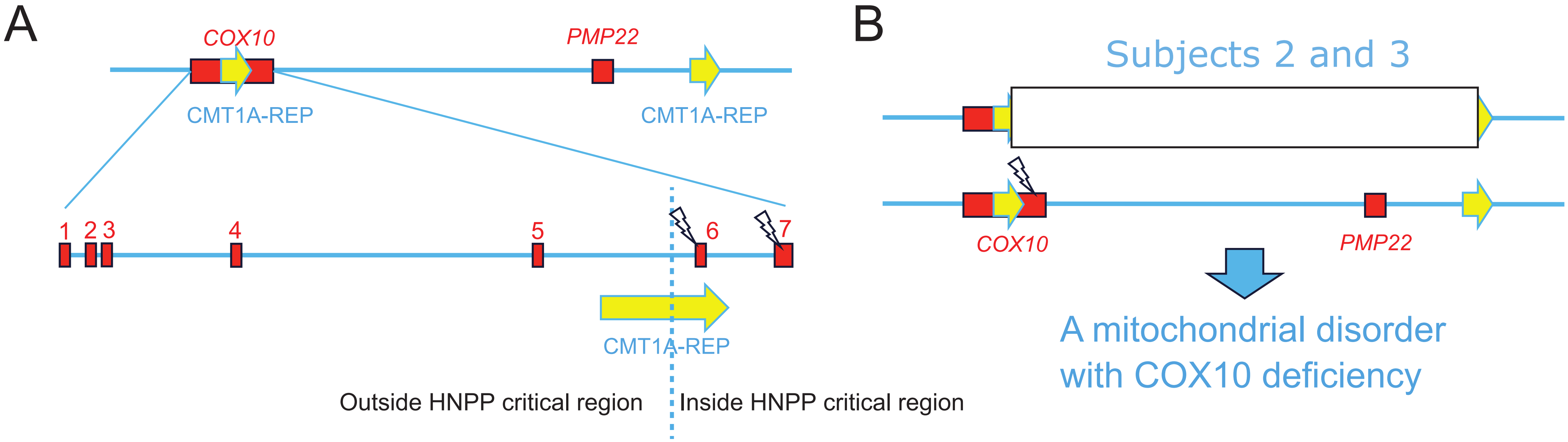
